# Large Language Models in Healthcare Simulation Education: A Bibliometric Analysis with AI-Assisted Screening

**DOI:** 10.64898/2026.06.02.26354722

**Authors:** Matthew Pears, Karan Wadhwa, Stephen R Payne, Stathis TH Konstantinidis, Chandra Shekhar Biyani

## Abstract

Large language models (LLMs) such as ChatGPT are rapidly reshaping healthcare education and simulation-based training in non-technical skills (NTS), yet no bibliometric analysis has mapped this landscape. We searched seven open-access databases (OpenAlex, PubMed, Europe PMC, Crossref, Semantic Scholar, CORE, DOAJ) for English-language publications from January 2020 to March 2026. From 100,277 initial records, a sequential keyword funnel yielded 830 candidate papers, which were screened by 83 independent Claude Sonnet 4.6 AI agents applying pre-specified inclusion criteria (PRISMA-trAIce compliant; Cohen’s kappa = 0.86 pre-reconciliation, 1.0 post-reconciliation). The final AI-verified corpus comprised 551 papers with a compound annual growth rate of 109%, contributions from 2,398 authors across 279 journals in 58 countries, and an h-index of 41. ChatGPT dominated the model landscape (46% of papers), with open-source models virtually absent. Virtual patient chatbots were the leading simulation modality (106 papers). Among NTS domains, communication (145 papers) and decision-making (135 papers) were most studied, whereas teamwork, leadership, situational awareness, and crisis resource management were markedly underrepresented. Only 6 urology-relevant papers were identified, none examining LLM integration within boot camp training formats. The field is growing at extraordinary pace but remains concentrated in a narrow range of NTS domains and a single proprietary model. Critical gaps persist in team-based skills training, open-source model evaluation, and specialty-specific simulation. AI-assisted bibliometric screening using multiple independent agents is feasible, reliable, and scalable, offering a replicable methodology for mapping fast-evolving research fields.

**Author Summary:** We mapped the research landscape of large language models in healthcare simulation and non-technical skills training by analysing 551 rigorously screened papers published between 2020 and 2026. Our analysis reveals a field that has exploded since the release of ChatGPT, growing at over 100% per year, but one with significant blind spots. Most research focuses on communication and clinical decision-making, while the team-based skills that prevent patient safety failures—teamwork, leadership, situational awareness, and crisis resource management—are barely studied. Almost half of all papers use a single proprietary model (ChatGPT), with open-source alternatives virtually absent. We also found that urology, despite having one of the most established simulation training programmes in surgery, has almost no research connecting large language models with simulation-based training. To conduct this analysis, we developed a novel screening approach using 83 independent AI agents, which achieved agreement with human review exceeding published benchmarks. Our open-access pipeline and dataset are freely available for other researchers to replicate or extend this work.

## 1. Introduction

The public release of ChatGPT in November 2022 triggered what is arguably the most rapid adoption of any technology in the history of healthcare education. Within months, researchers across medicine, nursing, and the allied health professions began exploring how large language models (LLMs), deep neural networks capable of generating, summarising, and reasoning over natural language, could be applied to clinical training, assessment, and decision support [1,2]. The subsequent literature has grown at a pace that challenges conventional methods of synthesis, demanding new approaches to mapping the research landscape, approaches that are themselves scalable, reproducible, and transparent.

Bibliometric analysis provides a systematic, quantitative framework for characterising scientific fields at scale [3]. Through established indicators such as Lotka’s law of author productivity, Bradford’s law of journal scattering, and Callon’s strategic thematic diagrams, bibliometric methods can reveal patterns of growth, collaboration, intellectual structure, and thematic concentration that narrative reviews cannot capture [4,5]. These methods are particularly well suited to emerging fields where the literature is too voluminous for traditional qualitative synthesis yet too immature for definitive systematic review. The intersection of LLMs and healthcare simulation is precisely such a field.

### 1.1 The ChatGPT Inflection Point

Healthcare simulation has been a cornerstone of medical education for over two decades, enabling learners to practise clinical skills in controlled, lower-risk environments across a continuum of modalities, from low-fidelity bench models and standardised patient encounters to high-fidelity mannequin scenarios and immersive virtual reality environments [6,7]. The advent of LLMs has introduced transformative possibilities across this continuum. Generative AI systems can simulate realistic patient interactions for communication training, generate scenario content for debriefing exercises, provide automated feedback on clinical reasoning, and serve as intelligent tutoring agents that adapt to individual learner performance [8]. Early demonstrations that GPT-4 could pass medical licensing examinations [9], and subsequent evidence that later models can generate clinically plausible case vignettes and role-play standardised patients with increasing fidelity [10], have prompted both considerable enthusiasm and legitimate concern regarding accuracy, bias, hallucination, and the pedagogical implications of machine-generated dialogue.

The scale of the resulting published research output is striking. From a starting pool of over 100,000 records across seven open-access databases, our screening pipeline identified 551 papers that substantively address LLMs in healthcare education and simulation—the majority published after 2022. Yet volume alone does not indicate maturity. Questions remain about which educational domains are being studied, which simulation modalities are being enhanced, which LLM platforms dominate the evidence base, and where critical gaps persist. These are the questions that bibliometric analysis is uniquely positioned to answer.

### 1.2 Non-Technical Skills: An Underexplored Frontier

While much early research on LLMs in healthcare has focused on knowledge retrieval and clinical reasoning, tasks closely aligned with technical competence, the role of these technologies in training non-technical skills (NTS) remains substantially less explored. Non-technical skills encompass the cognitive, social, and interpersonal competencies essential for safe clinical practice, including communication, teamwork, leadership, decision-making, situational awareness, and crisis resource management [11,12]. Failures in these competencies are implicated in a substantial proportion of adverse events across healthcare settings [11,13,14,15,16].

Simulation-based training is the predominant method for developing NTS in healthcare professionals. Structured programmes incorporating crisis resource management principles, standardised patient encounters, and facilitated debriefing have demonstrated measurable improvements in team performance and communication quality [17,18]. The potential for LLMs to augment this training is conceptually compelling: an AI-generated simulated patient could provide repeated opportunities for practising breaking bad news or managing difficult team dynamics. An LLM-powered debriefing assistant could offer structured post-scenario feedback aligned with established frameworks, such as the NOTSS. In urology specifically, Pears et al. conducted work on NTS training using ’bingo’-style cue identification techniques [19], evaluated a virtual reality-based NTS training application [20], and benchmarked a customised ChatGPT-4 model against consultant interaction for NTS feedback in simulated scenarios, finding that the AI demonstrated strengths in knowledge reinforcement, evidence-based feedback, and empathic communication [21].

Yet the distribution of research effort across NTS domains, simulation modalities, and clinical specialties remains unknown. Whether the literature is building a balanced evidence base or clustering around the most technically convenient applications, those that map most readily onto text-based interaction, is a question this analysis seeks to answer.

### 1.3 The Need for AI-Assisted Bibliometric Methods

A defining challenge when synthesising fast-moving research fields is the sheer volume of candidate literature. Traditional screening methods, in which human reviewers assess each record individually, become impractical when initial search yields exceed tens of thousands of records. This study addresses that challenge by deploying a hybrid screening methodology combining deterministic keyword filtering with AI-assisted verification using independent Claude Sonnet 4.6 agents, achieving inter-rater reliability exceeding the mean human-human kappa reported in the literature [22] (see Section 2.5 and 2.6 for full details). If AI-assisted screening can be shown to be reliable, transparent, and reproducible, it offers scalable infrastructure for living bibliometric analyses that can be re-executed at regular intervals as fields evolve, replacing the static snapshots that characterise traditional bibliometric studies with continuously updated evidence maps.

### 1.4 Rationale and Objectives

This study was motivated by three observations. First, research on LLMs in healthcare education and simulation is growing at a rate that exceeds the capacity of conventional review methods to remain current, yet while prior bibliometric studies have examined AI in medical education broadly [23,24], no published analysis has specifically mapped the scope, thematic distribution, and gaps in this literature. Second, the role of non-technical skills — communication, teamwork, leadership, decision-making, situational awareness, and crisis resource management — in simulation-based education is well established [11,12], but the extent to which LLM research engages with these competencies remains unknown. Understanding which NTS domains are being studied, and which are being neglected, is essential for guiding future research and curriculum development. Third, surgical specialties such as urology have well-established simulation training programmes [25] that appear well suited to LLM augmentation, yet the specialty-specific evidence base has not been characterised.

This analysis forms one component of a broader research programme from this papers’ authors examining the integration of LLMs into healthcare simulation. Companion studies include a Delphi consensus study on LLM implementation in simulation-based education, a scoping review of specific LLM applications, and a platform evaluation of an AI-powered simulation system. Together, these workstreams aim to provide an evidence base that is both methodologically rigorous and practically actionable for educators, simulation centres, and curriculum designers.

The specific objectives of this study are:

1. To map the publication landscape of LLMs in healthcare education and simulation from 2020 to 2026, characterising growth trajectories, geographic distribution, journal concentration, and model adoption patterns across a corpus of 551 papers.
2. To identify the dominant research themes, intellectual clusters, and emergent topics through keyword co-occurrence analysis and strategic thematic mapping.
3. To analyse the distribution of research across non-technical skills domains—determining which competencies (e.g., communication, decision-making) are well represented and which (e.g., teamwork, leadership, crisis resource management) remain comparatively underexplored.
4. To characterise the simulation modalities being studied in conjunction with LLMs, identifying where augmentation efforts are concentrated and where opportunities remain.
5. To quantify the representation of urology-specific research within the broader landscape and to identify gaps relevant to simulation boot camp training formats.
6. To demonstrate and validate a replicable, open-access, AI-assisted bibliometric pipeline that other researchers can execute without requiring institutional database subscriptions.

## 2. Methods

### 2.1 Study Design

This study employed a bibliometric design following established methodological frameworks [3,4]. The design was governed by two foundational principles: (a) analytical rigour consistent with contemporary bibliometric practice, and (b) full reproducibility using exclusively free, open-access data sources. The entire pipeline – data collection, deduplication, keyword filtering, AI-assisted screening, and bibliometric analysis – is publicly available on GitHub [26], enabling other researchers to replicate or update the study.

Reporting followed PRISMA 2020 guidelines [27], adapted for bibliometric analysis, and the PRISMA-trAIce checklist for transparent reporting of AI use in evidence synthesis [28]. The PRISMA-trAIce checklist items (M1-M9, R1-R2, D1-D2) are mapped to the corresponding sections throughout the Methods, with full details in Supplementary Appendix B.

### 2.2 Data Sources and Search Strategy

Seven electronic databases were systematically queried: OpenAlex, PubMed (via NCBI E-utilities), Europe PMC, Crossref, Semantic Scholar, CORE, and the Directory of Open Access Journals (DOAJ). These sources were selected for their complementary coverage of biomedical, clinical, and educational literature; free API access without institutional subscriptions; and sufficient metadata richness for bibliometric computation. The proprietary databases Web of Science and Scopus were deliberately excluded to maintain an open-access approach; the implications of this choice are considered in the limitations.

A total of 35 query strategies were executed across the seven sources. PubMed queries employed MeSH terms and title-abstract field tags. OpenAlex and Semantic Scholar used full-text and topic-level search filters where available. The remaining databases used keyword-based search adapted to their respective API capabilities. The complete query strings are provided in Supplementary Appendix A. The search was restricted to English-language publications from January 2020 to March 2026, with the start date chosen to capture a pre-ChatGPT baseline period within the timeframe of rapid LLM development.

### 2.3 Data Processing

Records from the seven databases were normalised to a common schema encompassing title, authors, year, DOI, abstract, journal/source, keywords, citation count, document type, and open-access status. Source-specific identifiers were retained for provenance tracking.

A three-stage deduplication algorithm was applied: exact DOI matching; fuzzy title matching (threshold of 90 on a 0-100 scale via the RapidFuzz library); and a fallback using author surname, publication year, and title-fragment matching. When duplicates were identified, a source-priority hierarchy (PubMed, OpenAlex, Europe PMC, Crossref, Semantic Scholar, CORE, DOAJ) determined which record was retained.

The initial search yielded **100,277** records. After deduplication, **86,635** unique records remained, corresponding to a deduplication rate of approximately 13.6%.

### 2.4 Screening Process: Sequential Keyword Funnel

Screening proceeded through a two-stage process: an automated sequential keyword funnel (Section 2.4) followed by AI-assisted verification of the filtered corpus (Section 2.5). This hybrid approach was designed to reduce a large initial corpus to a manageable set of candidate papers through deterministic keyword matching, before applying the more nuanced judgement of AI agents to distinguish genuinely relevant papers from false positives.

#### 2.4.1 Stage 1: Broad Keyword Funnel

The 86,635 unique records were filtered through three sequential keyword steps, each requiring the presence of domain-specific terminology in the title, abstract, or keywords:

- **Step 1 – LLM/AI filter:** Records were required to contain at least one term relating to large language models or generative AI (e.g., "large language model," "LLM," "ChatGPT," "GPT-4," "Claude," "Gemini," "generative AI," "foundation model"). This reduced the corpus from 86,635 to **37,280** records.
- **Step 2 – Healthcare/Medical/Education filter:** Remaining records were required to additionally contain at least one healthcare, medical, or education term (e.g., "healthcare," "clinical," "medical education," "nursing," "patient," "training"). This reduced the corpus to **29,238** records.
- **Step 3 – Simulation filter:** Remaining records were required to additionally contain at least one simulation-related term (e.g., "simulation," "virtual patient," "OSCE," "debriefing," "scenario," "mannequin," "VR," "XR"). This reduced the corpus to **5,725** records.

#### 2.4.2 Stage 2: Tighter Second-Pass Filter

A second-pass filter with stricter criteria was applied to the 5,725 records to further reduce noise before AI verification. This pass required: (a) a specific named LLM (e.g., "ChatGPT," "GPT-4," "Claude," "Gemini," "LLaMA," "Copilot") rather than generic AI terminology alone; (b) a medical or healthcare keyword in the title (not only the abstract); (c) the presence of at least one education-related keyword; and (d) exclusion of editorials and commentaries without original data. There were 830 papers remaining.

### 2.5 AI-Assisted Screening

#### 2.5.1 Screening Approach

AI-assisted screening was performed using Claude Sonnet 4.6 [29], accessed via the Anthropic API. The model was selected on the basis of published evidence demonstrating strong screening performance for LLM-based screening tools [30], including sensitivity of 0.81 across 23 Cochrane reviews [31], and accuracy of 0.945 approaching human-level performance (0.96) in obstetric literature screening [32].

AI was used at a single stage: verification of the 830 candidate papers identified by the keyword funnel, applying inclusion/exclusion criteria with the contextual judgement that keyword matching alone cannot provide. The 830 papers were distributed across 83 independent AI agents, each screening approximately 10 papers. Each agent received the title and abstract of each paper and was instructed to apply the study’s inclusion and exclusion criteria (Section 2.7), returning a binary include/exclude decision, confidence rating, tier classification (Tier 1–3), sub-analysis tags, and a brief rationale. No post-processing or override logic was applied prior to validation. The complete system prompt, per-paper instructions, and all raw agent outputs are available in the GitHub repository [26] and Supplementary Appendices B–D. The screening process complied with all applicable PRISMA-trAIce reporting items (M1–M9, R1–R2).

#### 2.5.2 Screening Outcomes and Corpus Composition

Of the 830 candidate papers screened by the AI agents, 551 (66.4%) were included and 279 (34%) were excluded. Of the 551 included papers, 80% of inclusion decisions were made with high confidence, with the remainder at medium or low confidence.

The included papers were classified into three tiers of increasing specificity: Tier 1 (LLMs in healthcare education broadly; n = 213, 38.7%), Tier 2 (LLMs in healthcare simulation; n = 193, 35%), and Tier 3 (LLMs in healthcare simulation with non-technical skills; n = 145, 26%). Sub-analysis tags were assigned as follows: 181 papers tagged as simulation-relevant, 78 as NTS-relevant, and 6 as urology-relevant. Open-access papers accounted for 297 of 551 records (54%).

Records in the final corpus were sourced from OpenAlex (n = 291), PubMed (n = 155), Europe PMC (n = 41), Crossref (n = 32), Semantic Scholar (n = 26), CORE (n = 6), and DOAJ (n = 0), reflecting the complementary coverage of the selected databases. The PRISMA flow diagram (Fig 1) details the complete selection process from the initial 100,277 records to the final 551-paper corpus.

**Fig 1.**
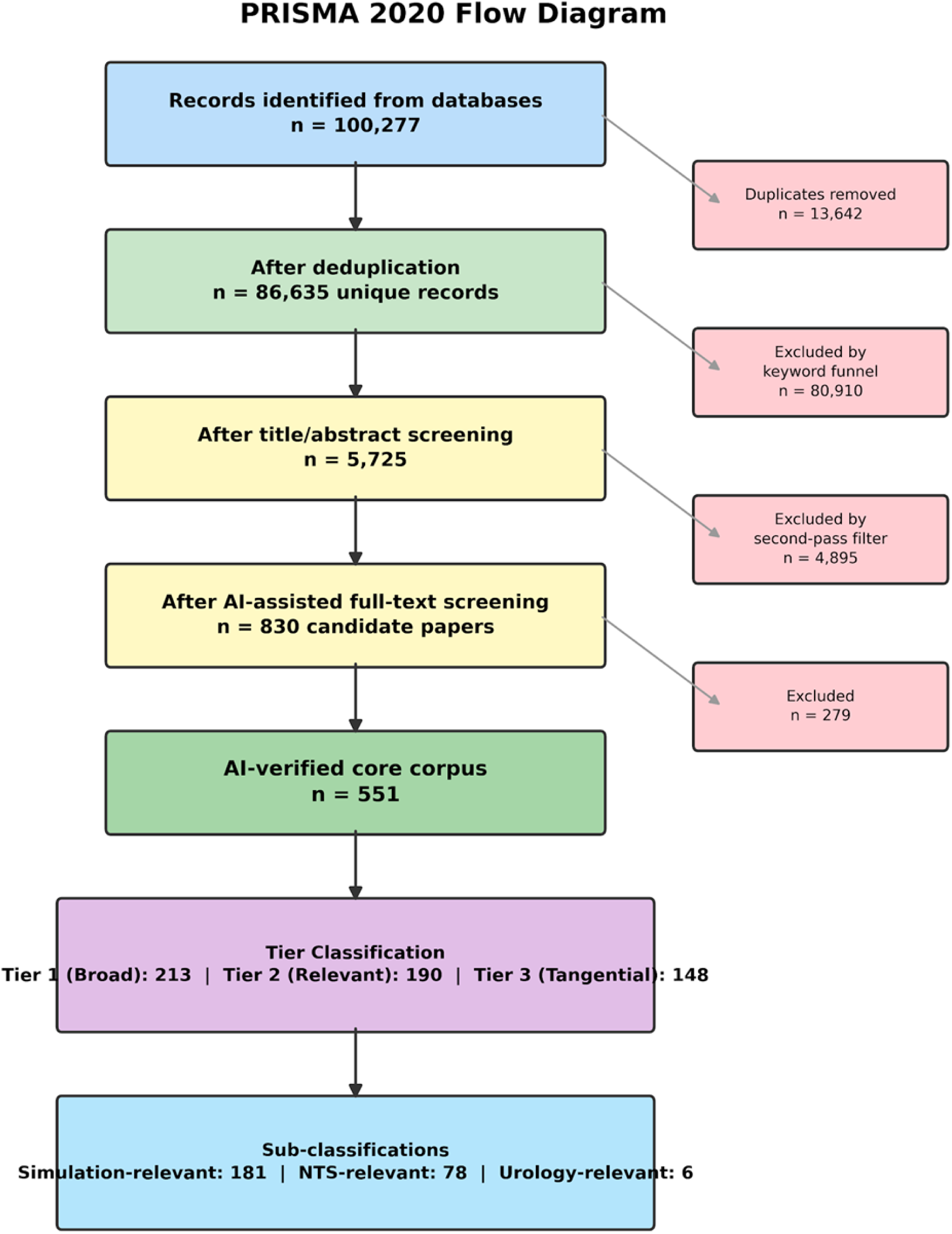
PRISMA 2020 flow diagram showing the study selection process.

### 2.6 Quality Assurance and Validation

AI-assisted screening was conducted in accordance with relevant position statements on AI use in evidence synthesis [33,34], and Table 1 summarises the methodological benchmarks used to evaluate screening quality.

| Benchmark | Threshold | This study | Interpretation |
| --- | --- | --- | --- |
| Cohen's kappa | 0.81-1.0 = almost perfect<br>0.61-0.8 = substantial | 0.9 pre-reconciliation<br>1.0 post-reconciliation | Agreement between raters beyond chance; 1.0 = perfect, 0 = random |
| Mean human-human kappa | 0.82 | Exceeded (0.9 and 1.0) | Average agreement between two human reviewers screening abstracts |
| Sensitivity | >0.9 desirable | 91.8% | Proportion of relevant papers correctly identified |
| Lotka's law exponent | Classical = 2 | 3.7 | Author productivity distribution; higher = fewer prolific authors |
| Bradford multiplier | Typically 3-5 | 5.4 | Journal concentration; higher = more concentrated |

The lead researcher (MP) independently reviewed 180 papers (22% of the 830-paper candidate corpus) across multiple validation stages. A formal 100-paper inter-rater reliability assessment yielded pre-reconciliation agreement of 94% (Cohen’s kappa = 0.86, a measure of agreement where values above 0.8 indicate "almost perfect" concordance), with all 6 disagreements involving exam benchmarking papers that lacked an educational intervention. Upon strict application of the pre-specified criteria, the reviewer agreed with the AI agents’ exclusion decisions, yielding full post-reconciliation agreement (Cohen’s kappa = 1.0). Both values exceed the mean human-human kappa of 0.82 for abstract screening reported by Hanegraaf et al. (2024) [22]. A review of all 830 agent-provided rationales identified zero errors of applying the desired criteria across all 83 agents. Human oversight was maintained throughout, with the lead author retaining final decision-making authority over all inclusion/exclusion decisions [34].

### 2.7 Inclusion and Exclusion Criteria

Papers were included if they met all of the following criteria:

a. addressed LLMs, generative AI, or a named system (e.g., ChatGPT, GPT-4, Claude, Gemini, other proprietary or open-source LLMs); b) were situated within a healthcare or clinical context (including medical, nursing, allied health, dental, and related domains); and c) were published between January 2020 and March 2026 in English.

Papers were additionally classified into tiers based on specificity:

- **Tier 1:** Met criteria (a)-(c) with a healthcare or medical education focus.
- **Tier 2:** Additionally required a simulation or training dimension (e.g., virtual patients, OSCEs, VR/AR/XR environments, scenario-based or mannequin-based simulation, debriefing).
- **Tier 3:** Additionally required explicit reference to at least one non-technical skills (NTS) domain (e.g., communication, teamwork, leadership, decision-making, situational awareness, crisis resource management).

Papers were excluded if they: addressed traditional machine learning without an LLM component; focused on clinical AI without an educational, training, or simulation dimension; addressed purely technical or computer science topics without a healthcare context; or were editorials, commentaries, or opinion pieces without original data or substantive methodological contribution. These criteria were applied consistently across both the keyword funnel and AI screening stages.

### 2.8 Bibliometric Analysis

All analyses were performed in Python using reproducible scripts. Unless otherwise indicated, all quantitative results in the Results section refer to the 551 papers in the final included corpus.

- Performance analysis included: annual publication trends with compound annual growth rate (CAGR); citation metrics (total, mean, median, h-index, g-index, i10-index); author productivity and collaboration patterns, including fit to Lotka’s law [35]; and journal concentration via Bradford’s law zones [36].
- Science mapping involved: keyword co-occurrence network construction (pairs co-occurring in ≥2 papers, individual keywords in ≥3 papers), and community detection via the Louvain algorithm [37].

**NTS domain classification** used regular-expression pattern matching across nine domains: decision-making, communication, teamwork, leadership, stress management, professionalism, situational awareness, task management, and crisis resource management (CRM). Both explicit terminology and closely related constructs (e.g., "empathy," "clinical reasoning," "shared mental model") were included in the patterns.

**Geographic analysis** extracted country-level authorship from affiliation strings where available. International collaborations were defined as multi-country authorship.

### 2.9 Sub-Analysis Strategy

Three sub-analyses were pre-specified to examine the literature at increasing levels of specificity:

- Simulation sub-analysis (n = 181): papers in which simulation-based education or training was a primary focus.
- Non-technical skills sub-analysis (n = 78): papers explicitly addressing one or more NTS domains in the context of LLM-assisted healthcare simulation.
- Urology sub-analysis (n = 6): the intersection of LLMs, simulation, and urological education, included to contextualise findings within a specific surgical specialty [21,19,20].

Each sub-analysis is presented separately in the Results.

## 3. Results

The screening pipeline described in Section 2 produced a final corpus of 551 papers classified into three tiers: Tier 1, LLMs in healthcare education broadly (n = 213, 39%); Tier 2, LLMs in healthcare simulation (n = 193, 35%); and Tier 3, LLMs in healthcare simulation with non-technical skills (n = 145, 26%). The complete selection process from the initial 100,277 records to the final corpus is presented in the PRISMA flow diagram (Fig 1). The following sections present the bibliometric characteristics of this corpus.

### 3.1 Publication Trends

The corpus exhibited explosive growth over the study period, with a compound annual growth rate (CAGR) of 109% (Table 1; Fig 3). Publication volume was almost entirely concentrated in the post-ChatGPT era: only four papers (0.7%) appeared before the release of ChatGPT in November 2022 (two in 2021 and two in 2022), whereas 547 papers (99%) were published from 2023 onward.

**Fig 2.**
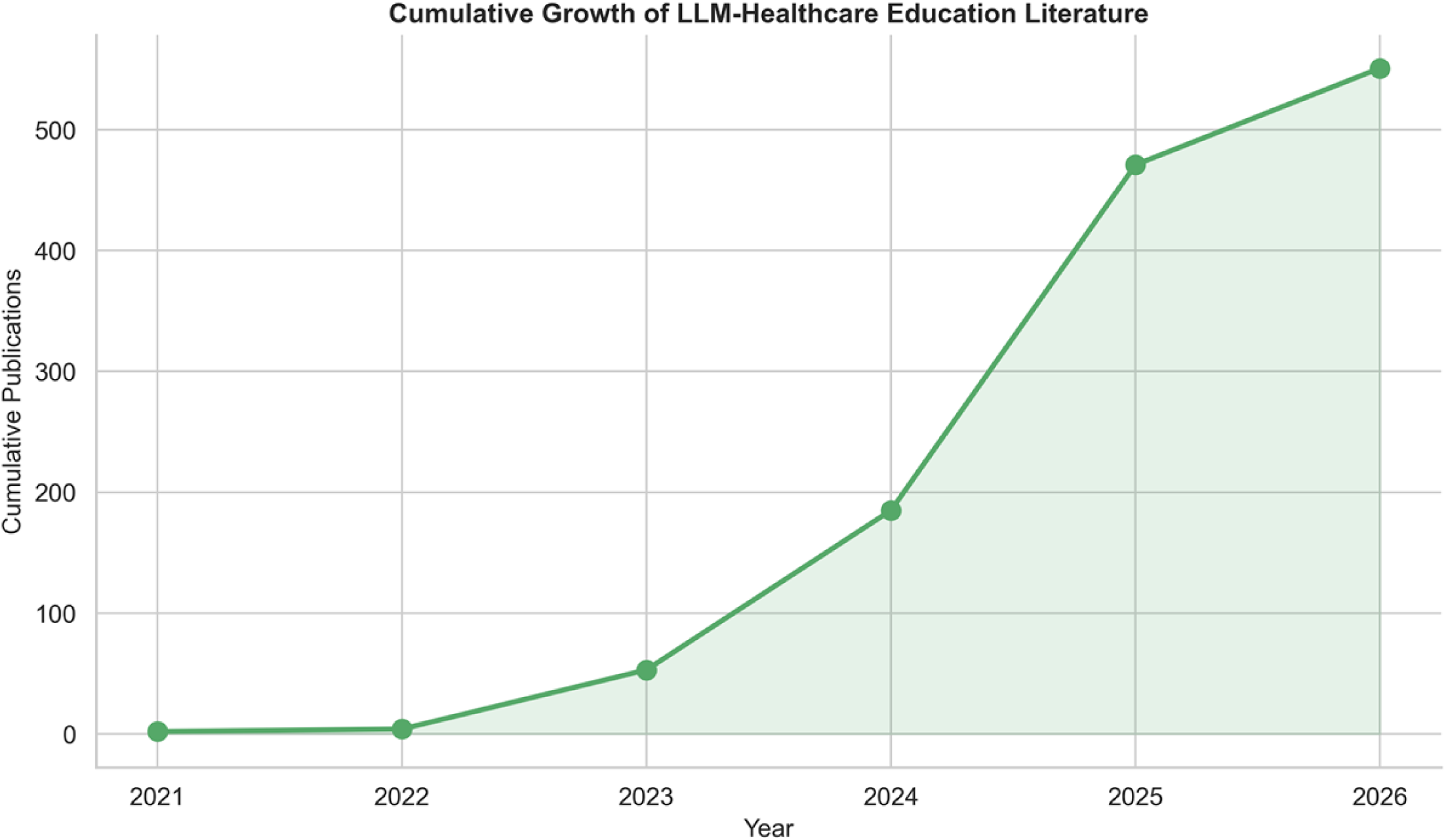
Cumulative publication growth over the study period.

**Fig 3.**
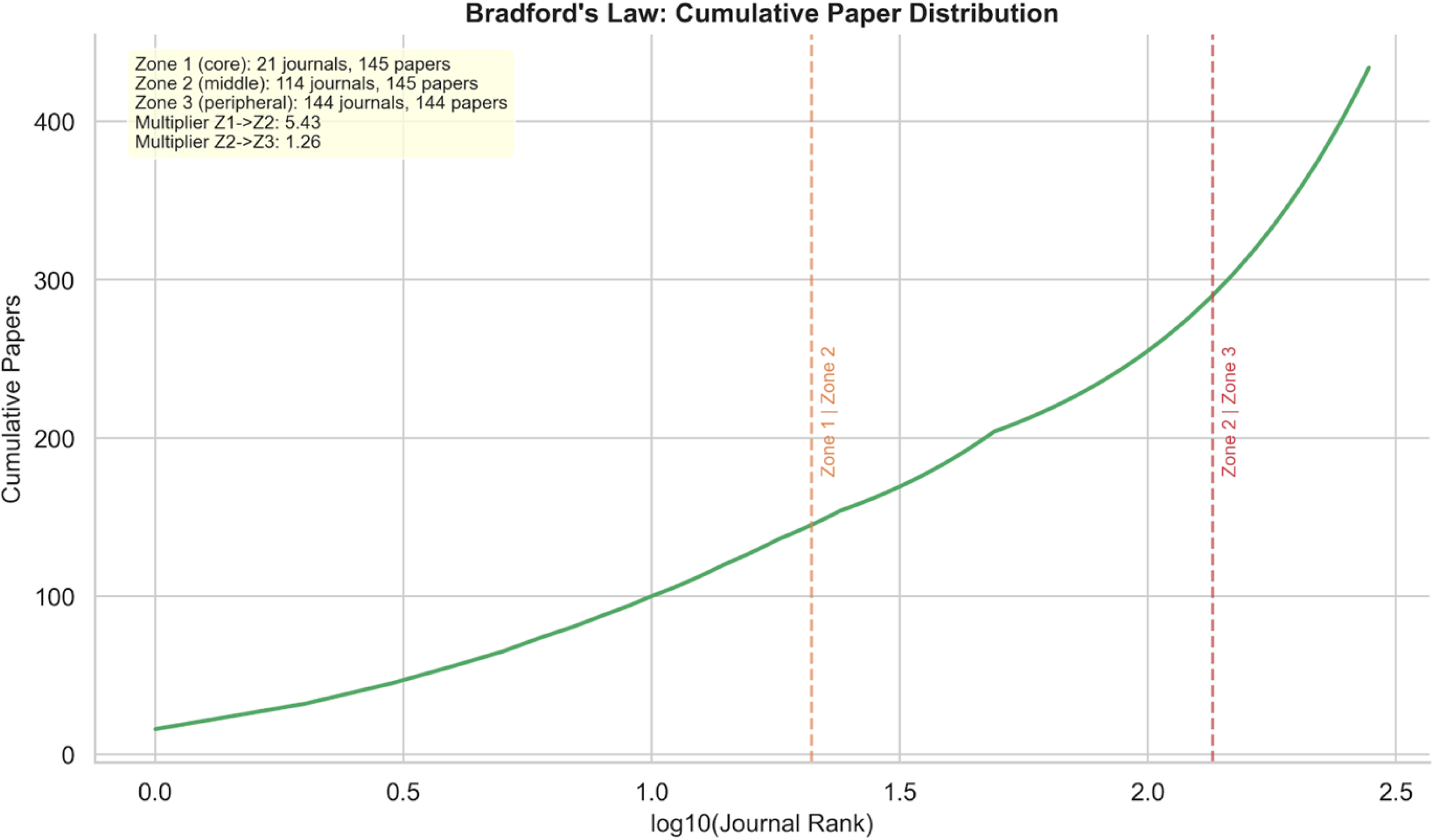
Bradford’s law journal zone analysis.

Annual output increased from 2 papers in 2021 and 2 in 2022 to 49 in 2023—a greater than 24-fold increase coinciding with the first full calendar year following ChatGPT’s release. Growth continued sharply: 132 papers in 2024 and 286 in 2025, the peak year, which alone accounted for 52% of all publications. Data for 2026 (n = 80 through March) indicate sustained high output; however, the partial-year figures preclude reliable full-year extrapolation.

The transition from 2022 to 2023 represents the largest inflection in the dataset, and the subsequent year-on-year increases (2.7-fold from 2023 to 2024, and 2.2-fold from 2024 to 2025) demonstrate that growth did not plateau after the initial surge. The median publication year was 2025, confirming that the literature is overwhelmingly recent. This growth trajectory is consistent with other bibliometric reports on generative AI in biomedicine and appears steeper than typical technology diffusion curves observed for many prior health technology innovations, suggesting a particularly rapid uptake of LLMs in healthcare education and simulation research.

### 3.2 Citation Analysis

The corpus accumulated a total of 6,448 citations, with a mean of 11.7 citations per paper and a median of 1.0 (Table 2). The citation distribution was heavily right-skewed: 222 papers (40%) were uncited at the time of data extraction, while a small number of highly cited works exerted disproportionate influence. The corpus h-index was 41 (41 papers with at least 41 citations each), the g-index was 69, and the i10-index was 131 (papers with 10 or more citations). The most cited paper received 550 citations.

Among the 20 most-cited papers (Table 3), the majority were published in 2023 and addressed foundational topics such as ChatGPT’s potential in medical education, its performance on clinical examinations, and opportunities and challenges for generative AI in health professions training.

The single most-cited work – "The rise of ChatGPT: Exploring its potential in medical education" (550 citations; *Anatomical Sciences Education*, 2023) – exemplifies the early exploratory phase of the field. Several highly cited papers specifically examined LLM-powered interactive medical simulations and simulated patient encounters, reflecting early recognition of the technology’s potential for experiential learning. Core journals for highly cited work included *JMIR Medical Education* (4 papers in the top 20), *BMC Medical Education*, *Cureus*, and specialty-specific outlets.

The high proportion of uncited papers (40%) alongside a relatively modest h-index of 41 reflects the extreme youth of this literature: the vast majority of papers were published in 2025 or later and have had limited time to accrue citations. Citation counts are therefore best interpreted as indicators of early influence rather than as measures of cumulative scholarly impact.

### 3.3 Author Analysis

A total of 2,398 unique authors contributed to the 551-paper corpus, yielding a collaboration index of 5.1 authors per paper (Table 4). This level of co-authorship is consistent with contemporary norms in biomedical and health informatics research and likely reflects the multidisciplinary expertise—spanning computer science, clinical practice, and education—required for LLM-related work.

Author productivity followed Lotka’s law with an exponent (beta) of 3.7 and an R-squared of 0.97, indicating an excellent fit. The Lotka exponent exceeds the classical inverse-square value of 2.0, indicating a steeper-than-expected productivity distribution in which prolific authors are even rarer than Lotka’s original formulation predicts. The majority of contributors (2,128 authors; 88.7%) published a single paper, while 270 authors (11.3%) contributed two or more. This high proportion of single-paper authors is characteristic of a young and rapidly expanding field in which many researchers are entering the domain rather than yet maintaining sustained, prolific research programmes.

The most productive authors were Li, S (9 papers), Li, Y and Wang, Y (7 papers each), and Wang, H (6 papers). However, the predominance of common East Asian surnames in the top-ranked positions warrants cautious interpretation, as name-based disambiguation across databases may conflate distinct individuals. Formal author disambiguation was beyond the scope of this study.

### 3.4 Journal Distribution

The 551 corpus papers spanned 279 unique journals or sources, with identifiable journal data available for the majority of records (Table 5; Fig 5). Bradford’s law analysis partitioned journals into three zones of approximately equal article yield (Table 6). Zone 1 (core) comprised 21 journals contributing 145 papers (34% of 434 papers with identifiable journal data), Zone 2 (middle) comprised 114 journals with 145 papers (33%), and Zone 3 (peripheral) comprised 144 journals with 144 papers (33%). The Bradford multiplier from Zone 1 to Zone 2 was 5.4, indicating a marked concentration of output among a small number of core outlets.

**Fig 4.**
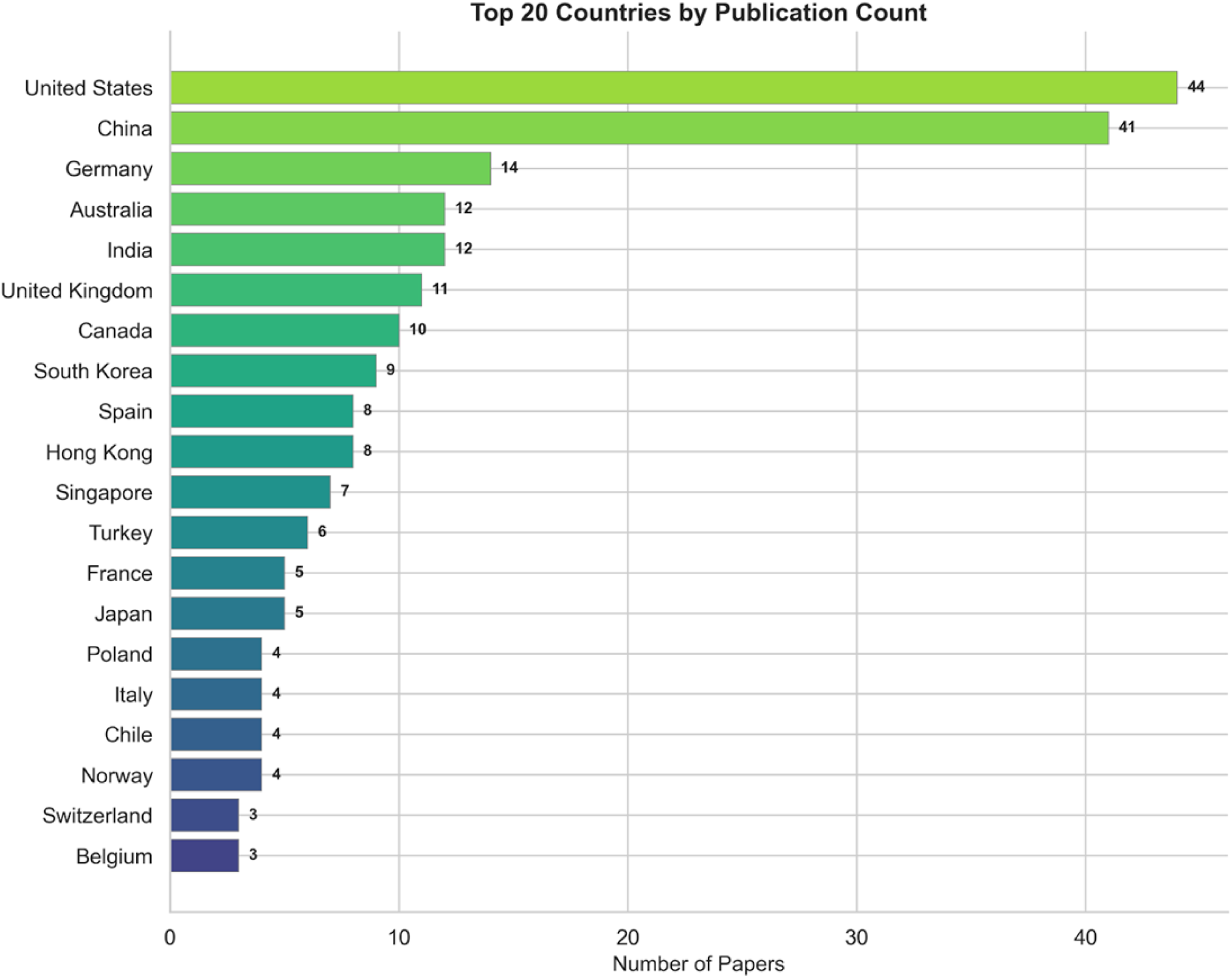
Geographic distribution of publications (top 20 countries).

**Fig 5.**
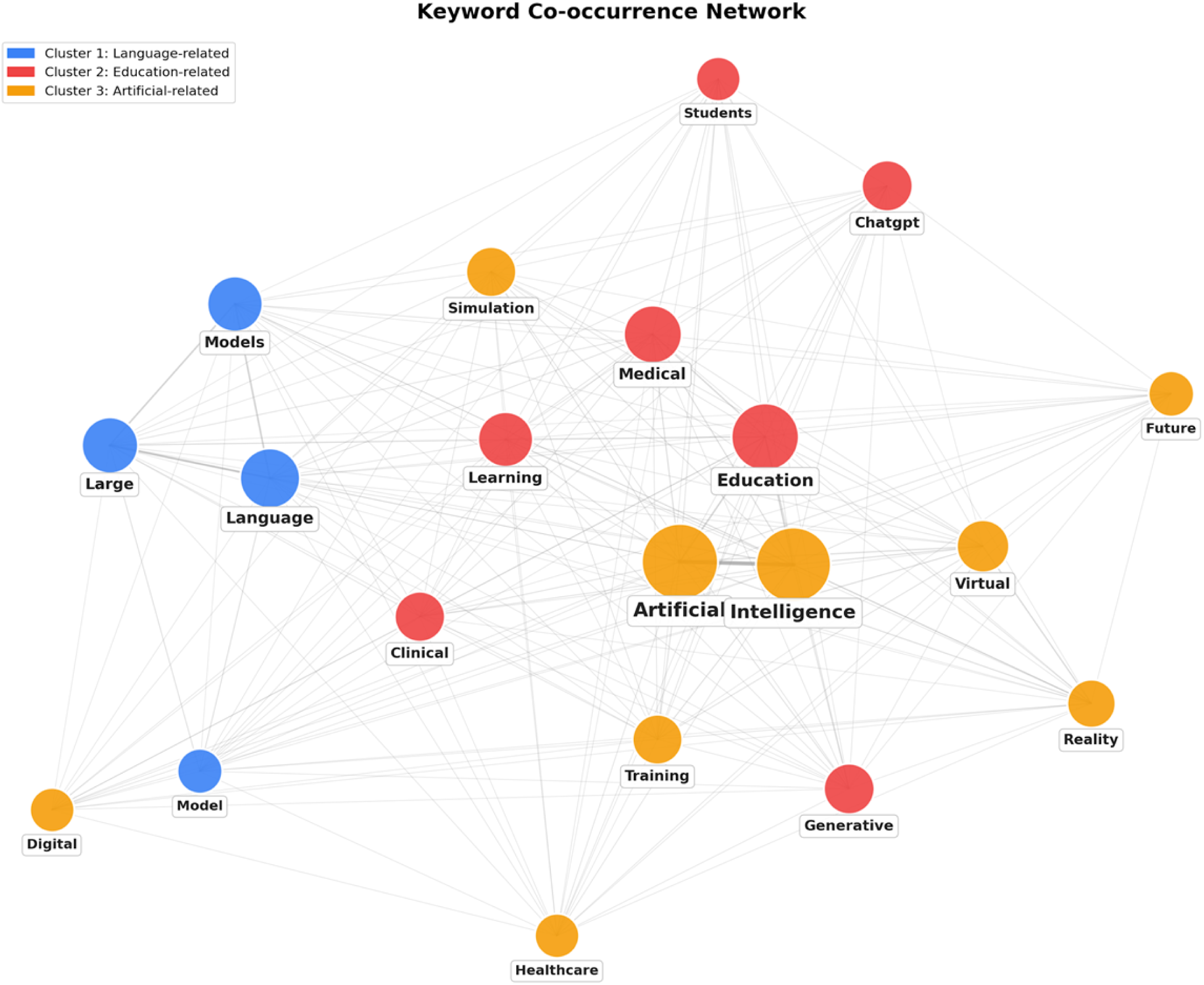
Keyword co-occurrence network with community clusters.

The three most prolific journals were *JMIR Medical Education* (16 papers; 3.7% when accounting for variant name entries), *BMC Medical Education* (16 papers; 3.7%), and *Cureus* (13 papers; 3%). Other prominent sources in the core zone included *Medical Teacher*, *arXiv* (as a preprint platform), *JMIR Formative Research*, *Clinical Simulation in Nursing*, and the *Journal of Medical Internet Research*. The presence of preprint repositories (arXiv, Open Science Framework, Preprints.org) among core sources is consistent with the field’s rapid dissemination norms and the broader open-science ethos in AI research.

The high degree of journal dispersion—with 144 peripheral-zone journals each contributing only 1 paper—underscores the interdisciplinary nature of the topic, which spans medical education, clinical specialties, health informatics, computer science, nursing education, and simulation science.

### 3.5 Geographic Distribution

Country-level authorship data were identifiable for 218 papers (39.6% of the corpus), revealing contributions from 58 countries (Table 7; Fig 6). The United States led in absolute output (44 papers; 20.2% of geographically identified papers), followed by China (41; 18.8%), Germany (14; 6.4%), Australia (12; 5.5%), and India (12; 5.5%). The United Kingdom (11 papers), Canada (10), South Korea (9), Spain (8), and Hong Kong (8) completed the top 10.

**Fig 6.**
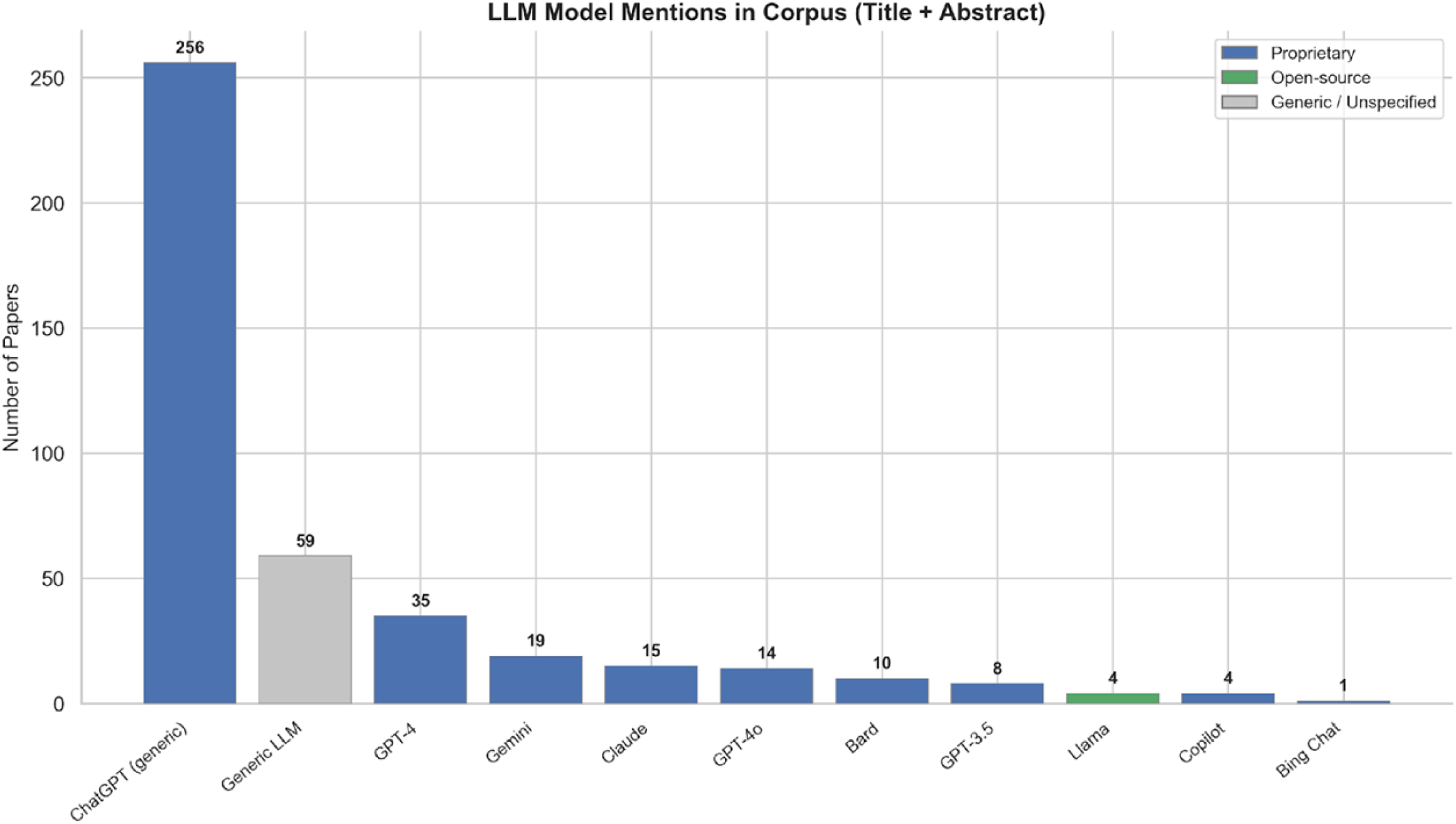
LLM model mentions frequencies across the corpus.

At the continental level, Asia accounted for the largest share of authorships (96; 34%), followed by Europe (79; 28.0%), North America (57; 20%), the Middle East (22; 7.8%), Oceania (14; 5%), South America (11; 3.9%), and Africa (3; 1.1%) (Fig 4). African (3; 1.1%) and South American (11; 3.9%) contributions were notably underrepresented.

International collaborations—defined as papers with authors from more than one country—accounted for 37 of the 218 geographically identified papers (17%), suggesting moderate but not dominant levels of cross-border cooperation. This may partly reflect the nature of much of this work, which frequently involves single-centre feasibility studies or evaluations that do not require multinational research teams.

### 3.6 Keyword Co-occurrence and Thematic Mapping

Keyword analysis of the corpus identified 6,412 unique keywords, with thematic clusters identified using co-word analysis methods [38]. The 10 most frequent keywords were "education" (appearing in 75.7% of papers), "clinical" (60.8%), "medical" (59.7%), "artificial" (57.7%), "intelligence" (57%), "language" (51.9%), "students" (44.1%), "learning" (43.7%), "large" (43.2%), and "chatgpt" (42.6%) (Table 8; Fig 8). The dominance of "education," "clinical," and "medical" as the top three terms confirms the corpus’s focus at the intersection of AI and health professions education.

**Fig 7.**
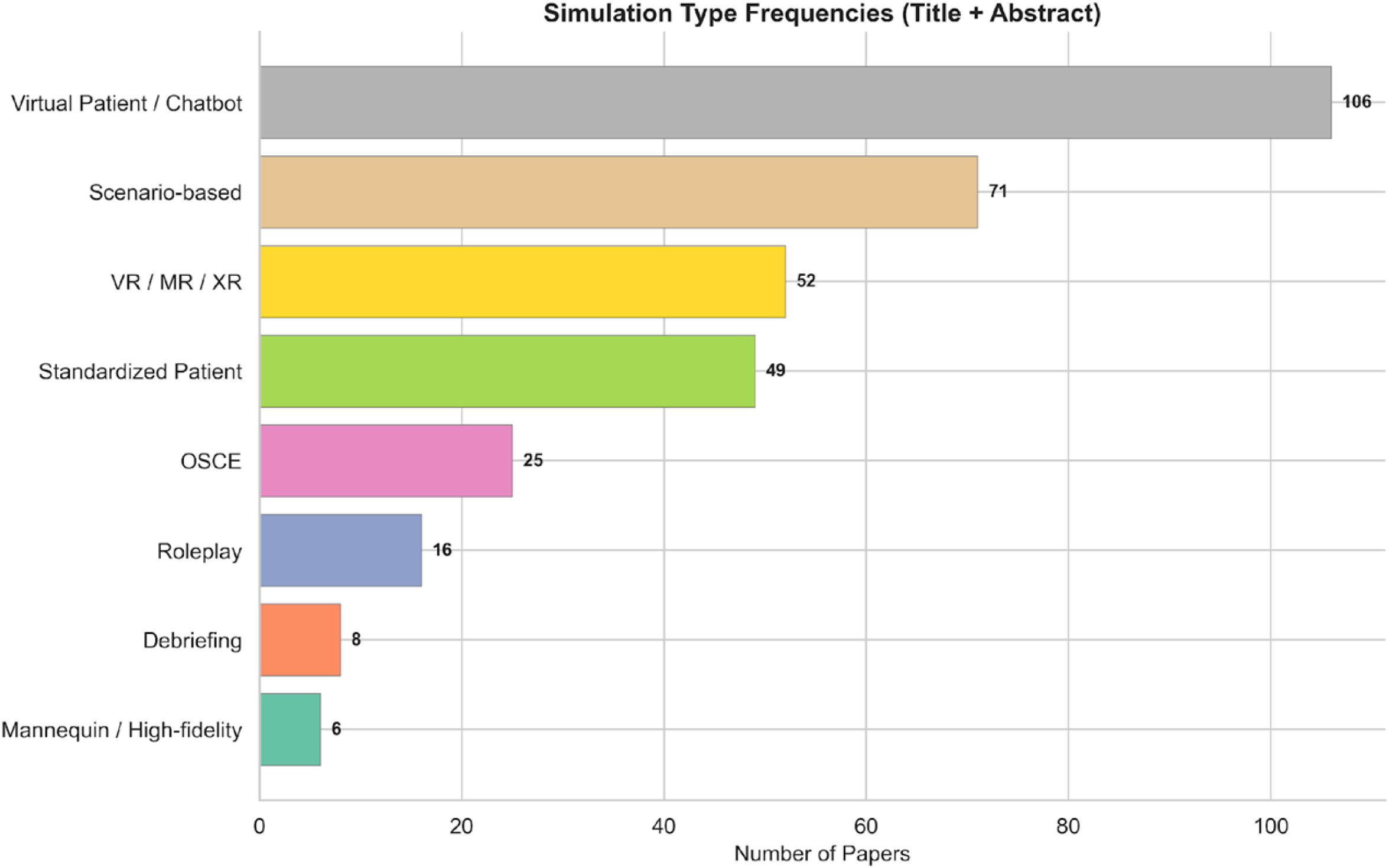
Simulation modality frequencies.

**Fig 8.**
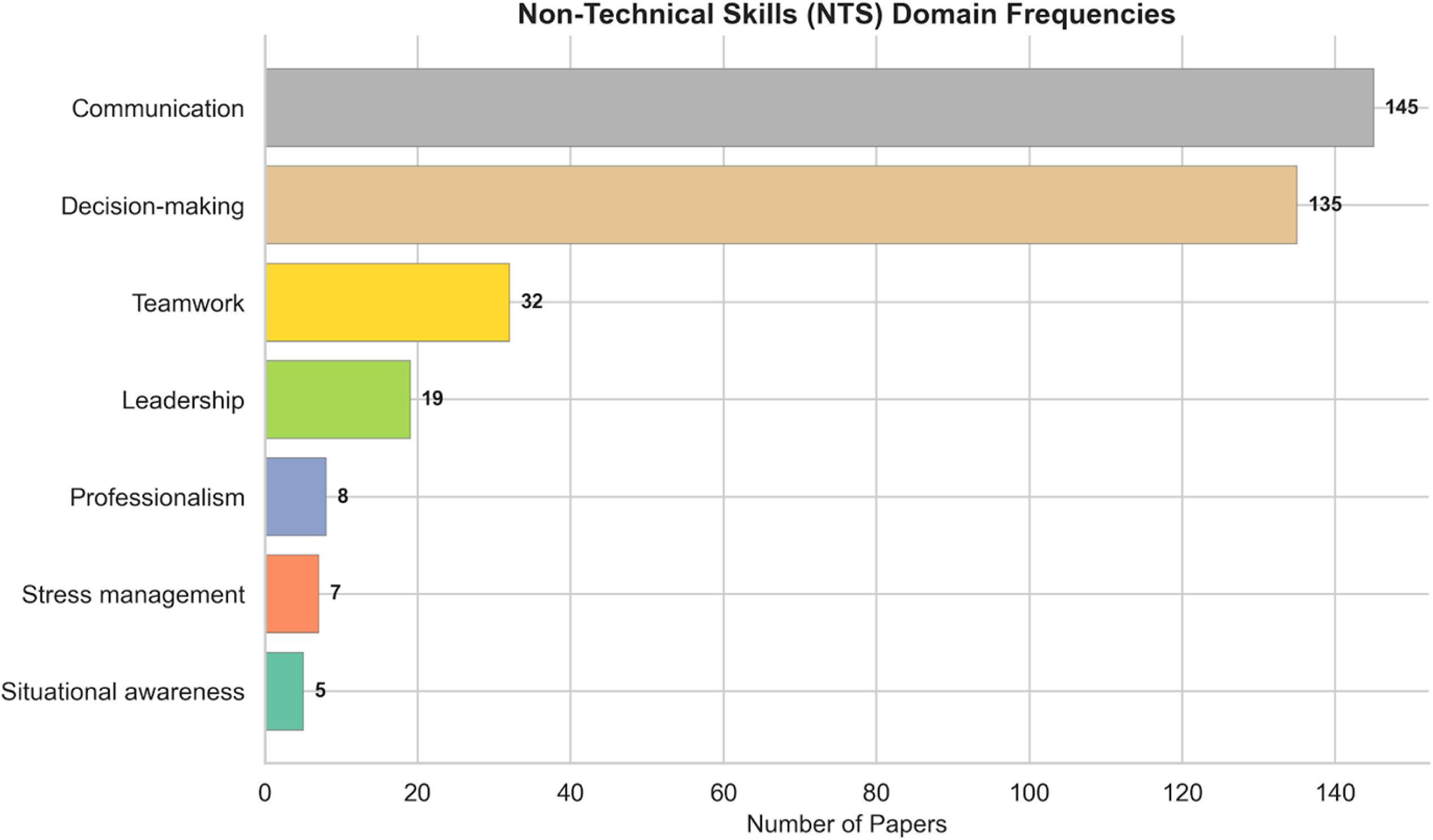
Non-technical skills domain frequencies.

The strongest keyword co-occurrence pairs were "artificial–intelligence" (313 co-occurrences), "education–medical" (267), "clinical–education" (259), and "language–large" (236), reflecting the core conceptual pillars of the field (Table 8). Notably, "simulation" appeared in 24.5% of papers and "feedback" in 20%, while "communication" (18.7%) and "assessment" (18.7%) featured prominently, consistent with the field’s orientation toward experiential learning and competency evaluation.

Keyword network analysis (Fig 5) and the associated word cloud further illustrate the thematic structure, with dense interconnections among AI/LLM terminology, educational concepts, and clinical domains. The co-occurrence of "virtual," "scenarios," and "patient" as high-frequency terms (28.9%, 27.9%, and 39.2%, respectively) points to the centrality of virtual patient and scenario-based applications in this literature.

### 3.7 LLM Model Landscape

Named-model classification revealed pronounced dominance by proprietary systems (Table 9). ChatGPT was by far the most frequently mentioned model, appearing in 256 papers (46.5% of the corpus). Generic references to "LLMs" without specifying a particular model appeared in 59 papers (10.7%). Among other explicitly named models, GPT-4 (35; 6.4%) and Gemini (19; 3.4%) were the next most common proprietary systems, followed by Claude (15; 2.7%), GPT-4o (14; 2.5%), Bard (10; 1.8%), GPT-3.5 (8; 1.5%), and Copilot (4; 0.7%).

Open-source models were almost entirely absent from the corpus. Llama appeared in only 4 papers (0.7%), and no other open-source model (e.g., Mistral) registered any mentions. Overall, 297 papers (54%) mentioned at least one proprietary model, whereas only 4 papers (0.7%) mentioned any open-source model. A negligible subset of 3 papers mentioned both proprietary and open-source models, indicating that direct comparative evaluation across licensing paradigms is essentially non-existent in this field.

Temporal trends in model mentions showed that ChatGPT has maintained dominant but stable representation across years. Gemini exhibited growth over time, while references to GPT-3.5 appear to be declining, consistent with model succession as newer versions superseded older ones.

### 3.8 Simulation Modalities

Simulation-type classification identified 266 of 551 papers (48.3%) as addressing at least one simulation modality (Table 10). Virtual Patient/Chatbot was the most prevalent modality (106 papers; 19% of the corpus), reflecting the natural alignment between conversational AI capabilities and interactive patient encounters. Scenario-based simulation was the second most common (71; 13%), followed by VR/MR/XR (52; 9.4%), Standardised Patient encounters (49; 8.9%), OSCEs (25; 4.5%), Roleplay (16; 2.9%), **Debriefing (8; 1.5%), and Mannequin/High-fidelity simulation (6; 1.1%).**

The prominence of Virtual Patient/Chatbot as the leading modality differs from the broader simulation literature, where mannequin-based and scenario-based approaches have historically predominated. This finding likely reflects the technological affordance of LLMs: their capacity for naturalistic dialogue generation maps most directly onto virtual patient and chatbot applications, making this the most intuitive entry point for LLM integration.

Mannequin-based simulation (6 papers; 1.1%) and debriefing-focused studies (8 papers; 1.5%) were notably underrepresented.

### 3.9 Non-Technical Skills Domains

Classification across non-technical skills (NTS) domains identified 282 of 551 papers (51.2%) as addressing at least one NTS domain, with 60 papers (11%) classified into multiple domains (Table 11). Communication was the most frequently addressed domain (145 papers; 26.3% of the corpus), followed closely by Decision-making (135; 24.5%). A substantial gap separated these two leading domains from the remaining skills: Teamwork appeared in 32 papers (5.8%), Leadership in 19 (3.4%), Professionalism in 8 (1.5%), Stress management in 7 (1.3%), and Situational awareness in 5 (0.9%).

Teamwork (32 papers), leadership (19 papers), stress management (7 papers), and situational awareness (5 papers) were markedly underrepresented relative to communication and decision-making.

Temporal trends in NTS domain representation showed growth across all domains, with Communication and Decision-making maintaining their leading positions throughout the study period. The absence of Crisis Resource Management (CRM) as a distinct category in the corpus is notable, given that CRM principles underpin the design and facilitation of many simulation scenarios in emergency medicine, anaesthesia, and surgery.

### 3.10 Urology Sub-Analysis

The urology-specific analysis identified 6 papers (1.1% of the corpus), indicating that the specialty has only just begun to engage with LLM-related research in education and simulation (Table 12). All 6 urology papers were published in 2024 or 2025. The subset accumulated 33 total citations (mean 5.5), and addressed topics including the feasibility of generative AI for history-taking in urology, AI-generated questions for urological competency assessment, non-technical skills training for urology trainees using ChatGPT-4, AI as a discriminator of competence in urological training, a chatbot for OSCE-based learning in nephrology/urology, and the use of ChatGPT for simulating Peyronie’s disease counselling.

NTS domains represented in the urology subset were Communication (2 papers) and Decision-making (1 paper). ChatGPT was the most frequently mentioned model (3 papers), with individual mentions of Gemini, GPT-4o, and GPT-4. The urology papers were evenly distributed across tiers (2 per tier), suggesting representation across the spectrum from broad healthcare education to simulation-specific and NTS-focused applications.

The intersection of LLMs, urology, and NTS-focused simulation training remains extremely sparse. Of the 6 identified papers, only the study by Pears et al. directly examined LLM use for NTS feedback in urology simulation, benchmarking ChatGPT-4 against consultant interaction for NTS feedback to urology trainees in simulated scenarios. No studies were identified evaluating LLMs within urology boot camp formats, which represent one of the most structured simulation training models in surgical education and would appear well suited to LLM augmentation – for example, through LLM-powered patient voices at mannequin stations, conversational virtual patients for history-taking practice, or AI-driven debriefing tools. The closest paper was Pears et al [21].

### 11 Tier Distribution and Corpus Structure

The three-tier classification provided a granular view of how the 551 papers relate to the study’s core focus (Table 13). Tier 1 papers (LLMs in healthcare education broadly; n = 213, 38.7%) formed the largest tier, reflecting the substantial volume of work on LLMs in medical education that does not specifically address simulation. Tier 2 papers (LLMs in healthcare simulation; n = 193, 35.0%) represented the simulation-focused subset, while Tier 3 papers (LLMs in healthcare simulation with NTS; n = 145, 26.3%) captured the most targeted literature at the intersection of all three domains.

Simulation-relevant flags were concentrated in Tiers 2 and 3 (110 and 69 papers, respectively), with only 2 Tier 1 papers flagged as simulation-relevant. NTS-relevant flags were predominantly assigned at Tier 3 (70 papers), with small numbers at Tier 1 (3 papers) and Tier 2 (5 papers). This distribution confirms the expected convergence of simulation and NTS themes in the most specific tier and validates the tiered classification as a meaningful stratification of the corpus.

Across tiers, growth was observed at all levels, with Tier 3 showing the steepest proportional increase in 2025 (89 papers, compared with 7 in 2023 and 28 in 2024), suggesting accelerating research interest specifically at the intersection of LLMs, simulation, and non-technical skills.

## 4. Discussion

### 4.1 Principal Findings

This bibliometric analysis mapped the research landscape of large language models in healthcare simulation and non-technical skills training from 2020 to 2026. The findings reveal a field that has emerged almost entirely since November 2022 and is expanding at an extraordinary pace, a compound annual growth rate of 109% with only four papers published before the release of ChatGPT and 547 published thereafter, yet one characterised by pronounced structural imbalances that have significant implications for research, education, and equity.

Five principal findings emerge. First, this is a field born of a single technological inflection: the release of ChatGPT. The near-total absence of publications before November 2022 (four papers in 2021–2022, compared with 286 in 2025 alone) distinguishes LLM-in-simulation research from most health technology adoption curves, which typically show gradual diffusion over years or decades [39]. This compressed timeline means the evidence base is being constructed in real time, with limited opportunity for the longitudinal validation that typically underpins educational innovation. Second, a single proprietary model dominates the research ecosystem: ChatGPT appeared in 46.5% of all papers, while open-source models were virtually absent (0.7% of papers). Third, virtual patients and chatbots have emerged as the dominant simulation modality (106 papers, 19%), signalling a fundamental shift in how standardised patient education may be delivered. Fourth, non-technical skills coverage is heavily skewed toward communication (145 papers) and decision-making (135 papers), with teamwork (32), leadership (19), and situational awareness (5) critically underrepresented. Fifth, the application of LLMs to simulation-based NTS training in urology is almost entirely absent from the literature, with only six papers identified in the entire corpus—a finding of direct relevance to the broader research programme of which this analysis forms a part.

### 4.2 The ChatGPT Inflection: What a 109.1% CAGR means for the field

The growth trajectory observed in this corpus is remarkable not merely for its steepness but for the sharpness of its onset. This speed carries both opportunity and risk. The opportunity is clear: rapid research output means the field is generating hypotheses, prototypes, and preliminary evidence at a pace that could accelerate the integration of LLMs into simulation curricula. The risk is equally clear: quantity does not guarantee quality. With 40.3% of the corpus remaining uncited at the time of data extraction and the median publication year being 2025, the vast majority of this literature has not yet been subjected to the scrutiny that comes with sustained citation, replication, and critical appraisal. The finding that 88.7% of authors (2,128 of 2,398) contributed only a single paper reinforces this concern; the field would benefit from sustained research programmes rather than isolated contributions.

Educators and institutions interpreting this literature should therefore exercise caution: the volume of publications should not be mistaken for the depth of evidence. Robust comparative effectiveness studies, particularly randomised controlled trials, remain scarce, and the rapid obsolescence of specific model versions (e.g., the decline of GPT-3.5 references from 8 papers across the corpus) means that findings tied to a particular model may have limited shelf life.

### 4.3 Model Concentration and the Open-Source Deficit

The most consequential structural finding is the extent to which the research ecosystem depends on a single commercial product. ChatGPT alone appeared in 46.5% of all papers (256 of 551). When other proprietary models are included, GPT-4 (6.4%), Gemini (3.4%), Claude (2.7%), GPT-4o (2.5%), proprietary systems collectively account for 53.9% of the corpus. Open-source models, by contrast, are almost entirely absent: Llama appeared in only four papers (0.7%), and no other open-source model exceeded this figure. Only three papers in the entire corpus mentioned both proprietary and open-source models, indicating that systematic comparative evaluation across licensing paradigms is virtually non-existent.

This concentration raises concerns regarding reproducibility (proprietary models are opaque and subject to unannounced updates), accessibility (commercial pricing creates barriers for low-resource institutions), and generalisability (findings from a single model may not transfer to other architectures).

### 4.4 Virtual Patients as the Dominant Simulation Paradigm

The emergence of virtual patients and chatbots as the leading simulation modality in this corpus (106 papers, 19%) represents a significant finding. This modality surpassed scenario-based simulation (71 papers, 13%), VR/MR/XR (52 papers, 9.4%), and standardised patient encounters (49 papers, 9%). The prominence of virtual patients likely reflects the natural alignment between conversational LLMs and the core educational task of patient interaction: LLMs are, at their most fundamental level, dialogue systems, and simulated patient conversations represent an application domain where the technology’s strengths, fluent natural language generation, contextual responsiveness, and scalability, directly serve the educational purpose.

This finding has significant implications for standardised patient (SP) programmes, a cornerstone of medical education worldwide. Traditional SP encounters are resource-intensive, requiring trained actors, dedicated scheduling, and physical infrastructure. LLM-powered virtual patients offer the prospect of unlimited practice opportunities, immediate availability, consistent performance, multilingual capability, and automated assessment, advantages that could fundamentally alter the economics and accessibility of communication skills training. However, the educational equivalence of virtual patient encounters to human SP interactions has yet to be established through rigorous comparative studies. Critical questions remain about whether LLM-generated patient personas can replicate the emotional complexity, non-verbal communication, and unpredictability that characterise real patient encounters, and whether learners develop transferable skills from interactions with AI rather than human interlocutors.

The relatively low representation of mannequin-based and high-fidelity simulation (6 papers, 1.1%) is notable given that this modality is arguably the most amenable to LLM augmentation through text-to-speech interfaces. Mannequin-based scenarios in which human operators currently provide simulated patient voices could be enhanced or partially automated through LLM-driven conversational agents, potentially improving consistency and scalability. The near-absence of this application in the literature suggests an underexplored opportunity.

### 4.5 The Non-Technical Skills Gap

The distribution of NTS research across classified domains reveals a hierarchy that appears to reflect technological convenience rather than clinical importance. Communication (145 papers, 26.3%) and decision-making (135 papers, 24.5%) together account for the majority of NTS-related research, likely because these competencies align most naturally with the text-based capabilities of current LLMs: clinical reasoning maps onto question-answering paradigms, and communication training maps onto conversational interaction. Together, these two domains generated 280 of all NTS domain-level classifications; given that 60 papers were classified into multiple domains, the true number of unique papers addressing communication or decision-making is likely somewhat lower, but the concentration is nonetheless striking.

Critically, the decision-making studied in the current literature is predominantly individual clinical reasoning in controlled settings, not the dynamic, time-pressured decision-making that occurs during clinical crises. Crisis resource management (CRM) requires the simultaneous deployment of multiple NTS domains—leadership, teamwork, situational awareness, and communication—under conditions of stress, uncertainty, and evolving clinical deterioration.

These are fundamentally different cognitive and interpersonal demands from those tested by an LLM presenting a diagnostic vignette. The near-absence of CRM, stress management, and situational awareness from the literature therefore represents not merely a quantitative gap but a qualitative one: the NTS domains most critical to patient safety in high-acuity settings remain almost entirely unaddressed by LLM research.

The deficits at the other end of the spectrum are stark. Teamwork appeared in only 32 papers (5.8%), leadership in 19 (3.4%), professionalism in 8 (1.5%), stress management in 7 (1.3%), and situational awareness in 5 (0.9%). These are not peripheral competencies; they are central to patient safety and have been the focus of decades of human factors and simulation-based training research [11,12,40,41].

The near-absence of situational awareness from the LLM literature is particularly striking given its foundational role in Endsley’s model of decision-making in dynamic systems [40] and its centrality to crisis management training in anaesthesia, emergency medicine, and surgery. The very low representation of crisis resource management (CRM) principles—despite CRM being foundational to high-quality simulation design, since its adaptation from aviation by Gaba and colleagues [41]—further underscores this imbalance.

The disparity cannot be explained by the relative importance of these competencies to clinical practice. Rather, it likely reflects two structural factors. First, communication and decision-making are inherently individual, text-mediated competencies that align with the current architecture of single-agent LLM systems. Teamwork, leadership, and CRM, by contrast, are inherently team-based, dynamic, and context-dependent, qualities that require multi-agent interaction, real-time coordination, and environmental awareness that current LLM architectures are poorly equipped to address in isolation.

Second, the dominance of virtual patient and chatbot modalities (which naturally emphasise one-to-one conversation) may further channel research toward communication at the expense of team-based competencies. This gap has practical implications. Simulation programmes that integrate LLMs for communication or clinical reasoning training may inadvertently neglect the team-based, systems-level competencies that are most strongly associated with adverse event prevention.

Future research should explicitly address how LLMs might support these underrepresented domains—for example, through multi-agent systems that simulate team dynamics, through intelligent debriefing tools that analyse team communication patterns against CRM frameworks, or through integration with physiological monitoring systems to create realistic stress and situational awareness challenges.

### 4.6 The Urology Gap: A Major Finding for Specialty-Specific Simulation

The urology sub-analysis represents one of the most significant findings of this study in the context of the broader research programme. Of the 551 papers in the corpus, only six (1.1%) addressed the intersection of LLMs and urology. Within this subset, only two papers were classified under communication and one under decision-making; no papers addressed teamwork, leadership, situational awareness, or any other NTS domain in the urological context.

#### This finding is noteworthy for several reasons

First, urology has one of the most established simulation training infrastructures of any surgical specialty. The UK urology boot camp programme, now in its tenth year and recommended by the Joint Committee on Surgical Training for all new registrars, represents a structured, well-resourced, and widely adopted simulation format [25].

These boot camps typically employ mannequin-based stations with human operators providing simulated patient voices, standardised patient encounters for communication training, and facilitated debriefing sessions—each of which is amenable to LLM augmentation. Yet no studies in the corpus evaluated LLMs within boot camp formats.

Second, the near-absence of LLM-NTS research in urology exists despite emerging evidence that the technology can meaningfully contribute to NTS development in the specialty. The work of Pears et al. [21] demonstrated in a double-blinded study that a customised ChatGPT-4 exhibited significant strengths in reinforcing knowledge gathering, providing evidence-based feedback, conveying empathy, and tailoring explanations to complexity when benchmarked against consultant interaction for NTS feedback to urology trainees. This suggests that the technology is viable; the gap lies in the absence of further research building on these findings.

Third, the fact that only six papers specifically address urology in a simulation context, despite the growing volume of urology-related LLM research more broadly, illustrates the broader pattern observed across the corpus: the majority of LLM research in clinical specialties focuses on examination performance, patient information quality, and clinical decision support rather than on simulation-based NTS training. This represents an opportunity for research groups positioned at the intersection of urology, simulation, and AI.

Emerging platforms are beginning to address this gap. The Simuro platform (simuro.net), developed as part of the present research programme, is among the first purpose-built LLM-powered simulation systems designed specifically for surgical specialty training, integrating conversational AI with NTS-focused scenarios for urology education. A companion Delphi consensus study (in preparation) aims to establish governance frameworks for AI implementation in simulation-based education. Together, these workstreams seek to bridge the gap between the technology’s demonstrated potential and its current near-absence from the urology simulation evidence base.

### 4.7 Geographic Concentration and Global Equity

Country-level authorship data were available for 218 papers (40% of the corpus), revealing contributions from 58 countries. The United States (44 papers) and China (41 papers) together accounted for 39% of geographically identified papers, followed by Germany (14), Australia (12), India (12), and the United Kingdom (11). At the continental level, Asia (34%), Europe (28%), and North America (20%) collectively accounted for 82% of identified authorships, while Africa contributed only three papers (1.1%).

This concentration raises concerns about whose perspectives and contexts shape the emerging evidence base for LLM integration in healthcare simulation. If the majority of research is conducted in well-resourced institutions in high-income countries, the resulting tools, curricula, and evaluation frameworks may not be appropriate for or transferable to the settings where scalable AI-powered training could have the greatest impact. Low- and middle-income countries often face the most acute workforce shortages and the greatest need for scalable, cost-effective training solutions, yet they are precisely the regions most underrepresented in the research and most likely to face barriers to accessing proprietary LLM platforms.

### 4.8 AI-Assisted Screening: Methodological Implications (PRISMA-trAIce D1, D2)

This study employed a hybrid screening methodology combining deterministic keyword filtering with AI-assisted verification using Claude Sonnet 4.6, reported in accordance with the PRISMA-trAIce checklist [28]. The approach reduced 100,277 initial records to 551 included papers through a transparent, reproducible pipeline, with all code, data, and AI prompts hosted in a public GitHub repository to enable replication regardless of institutional subscriptions.

The achievement of a Cohen’s kappa of 0.86 pre-reconciliation (1.0 post-reconciliation) between AI and human reviewer—exceeding the mean human-human kappa of 0.82 reported in the literature [22]—provides empirical support for LLMs as screening tools in bibliometric research. However, AI screening introduces qualitatively different errors from human review: LLM-based screeners may exhibit systematic biases reflecting their training data, and residual misclassification in unsampled papers cannot be excluded.

AI-assisted screening is now a practical reality for evidence synthesis. The Cochrane, Campbell, JBI, and CEE position statements [33,34] endorse AI as a scalable screening tool provided that transparency and human authority are maintained, and the PRISMA-trAIce framework [28] provides the reporting infrastructure to support this. As these methods mature, they have the potential to make large-scale bibliometric analysis accessible to research teams without the resources for traditional multi-reviewer screening.

### 4.9 Limitations

Several limitations should be considered when interpreting these results. First, the exclusion of Scopus and Web of Science means that publications indexed exclusively in these proprietary databases may have been missed. However, the growing overlap between OpenAlex and these sources [42], combined with seven complementary databases, mitigates this concern. The 551-paper corpus represents the most rigorously screened subset—each paper individually verified by AI agents and audited by the lead author.

Second, AI-assisted screening, while validated against human reviewers with a Cohen’s kappa of 0.86 pre-reconciliation and compliant with PRISMA-trAIce guidelines, may introduce classification errors distinct from those of human reviewers. In particular, title-and-abstract-only screening—without access to full text—means that papers whose relevance is apparent only from their methods or results sections may have been misclassified. The validation protocol assessed a stratified sample; residual misclassification in the unsampled papers cannot be excluded.

Third, the screening pipeline used a single LLM (Claude Sonnet 4.6) for AI-assisted verification. While this model was selected on the basis of published validation evidence [31,32], using the same class of technology (LLMs) to screen a literature about LLMs introduces a potential reflexive bias: the screening model may have differential sensitivity to papers about models from its own family or from competing commercial platforms. Future studies could mitigate this by employing multiple LLMs for screening and comparing their inclusion decisions.

Fourth, NTS domain classification relies on keyword-based regular expression matching, which captures explicit terminology but may miss papers addressing NTS concepts without using standard vocabulary. This approach likely underestimates the true prevalence of NTS research, particularly for domains such as situational awareness that may be discussed using non-standard language. Conversely, broad terms such as "leadership" may capture papers addressing organisational rather than clinical NTS, potentially overestimating certain domains.

Fifth, country-level geographic data were available for only 39.6% of the corpus, limiting the generalisability of geographic findings. The concentration patterns observed may not fully represent the global distribution of research activity.

Sixth, citation data represent a snapshot at the time of extraction (March 2026) and will change as the literature matures. The 40% uncited rate largely reflects the fact that 52% of papers were published in 2025 and 14.5% in 2026 (through March), leaving insufficient time for citation accumulation. Citation-based indicators should be interpreted with this temporal constraint in mind.

Seventh, the single-reviewer design (MP) for human validation, while augmented by AI screening and consistent with the resource constraints of the study, means that the human component of the quality assurance process lacked independent dual-reviewer verification. The high AI-human agreement (Cohen’s kappa = 0.86 pre-reconciliation, 1.0 post-reconciliation) partially compensates for this limitation but does not fully replace dual human review.

Finally, the corpus of 551 papers represents the AI-verified subset of a larger literature; the keyword-based filtering stages preceding AI verification may have excluded relevant papers that did not contain the specified terminology. The trade-off between sensitivity and specificity in the filtering pipeline is inherent to the approach and is documented transparently in the methods.

### 4.10 Implications for Practice

The findings of this analysis carry several implications for educators, researchers, and institutions. For **simulation educators**, the data indicate that LLM applications are currently most mature for communication and decision-making training, particularly through virtual patient and chatbot modalities. Educators seeking to integrate LLMs should begin with these applications while recognising the need to maintain human-led facilitation for teamwork, CRM, and other team-based competencies where AI capabilities remain limited. The virtual patient modality appears particularly promising as a scalable supplement to—though not a replacement for—standardised patient programmes.

For **researchers**, the analysis identifies several priority areas: the critical underrepresentation of situational awareness, stress management, and CRM in the LLM literature; the near-total absence of open-source model evaluation; the unexplored potential of LLM augmentation for mannequin-based and high-fidelity simulation; and the need for specialty-specific studies in disciplines such as urology where structured simulation programmes already exist. The finding that 89% of authors contributed only a single paper suggests that many researchers are conducting exploratory studies rather than sustained programmes of inquiry; the field would benefit from dedicated research groups pursuing longitudinal agendas.

For **institutions** integrating LLMs into simulation programmes, the findings highlight specific opportunities and risks. The critical gaps in teamwork, leadership, situational awareness, and CRM-focused training represent priority areas where purpose-built simulation platforms could have the greatest educational impact—particularly in procedural specialties such as urology, where established boot camp and NTS training infrastructures already exist but LLM integration remains virtually absent. However, translating these opportunities into practice requires substantial investment: purpose-built simulation platforms demand specialist software development, careful pedagogical design, rigorous usability testing, and seamless integration with existing curricula—resources that few simulation centres currently possess. Institutions pursuing LLM-powered simulation should plan for these development timelines and consider collaborative models that share technical expertise across training programmes.

### 4.11 Future Directions

The landscape mapped in this analysis points to several priority areas for future research.

First, the field urgently requires **randomised controlled trials** examining the educational effectiveness of LLM-integrated simulation compared with traditional approaches. The evidence base is dominated by descriptive, development, and feasibility studies; without rigorous comparative evidence, the educational value proposition of LLMs in simulation remains theoretical.

Second, the NTS gaps identified here—particularly in **situational awareness, stress management, teamwork, and leadership**—demand targeted investigation. Multi-agent LLM systems that simulate team dynamics, real-time physiological monitoring integrated with LLM-driven scenarios, and intelligent debriefing tools that analyse team performance against established NTS frameworks represent promising research directions that could address these deficits.

Third, the **urology simulation gap** warrants prospective studies that directly evaluate LLM integration within established training formats. The UK urology boot camp programme [25] offers a particularly suitable testbed: LLM-powered patient voices at mannequin stations, conversational virtual patients for history-taking and breaking-bad-news practice, and AI-driven debriefing tools aligned with the NoTSUS framework represent feasible interventions that could be evaluated within existing curricular structures. The Simuro platform, developed as part of this research programme, provides a purpose-built infrastructure for such evaluation. More broadly, other procedural specialties with established simulation programmes—orthopaedics, neurosurgery, vascular surgery—would benefit from analogous specialty-specific investigations.

Fourth, the **open-access bibliometric pipeline** developed for this analysis should be leveraged for longitudinal monitoring of the field. Given the 109% compound annual growth rate, the landscape described here will evolve substantially within months. Planned re-execution of the pipeline at six-month intervals would create a living bibliometric resource, enabling the research community to track the evolution of themes, model adoption, NTS coverage, and geographic distribution in near-real time. Such living bibliometric analyses could replace the static snapshots that characterise traditional bibliometric studies with continuously updated evidence maps.

## 5. Conclusions

This bibliometric analysis of 551 AI-verified papers, drawn from 100,277 initial records across seven open-access databases, provides the first comprehensive map of research on large language models in healthcare simulation and non-technical skills training from 2020 to 2026. Six principal conclusions emerge.

First, the field has grown at an extraordinary pace (109% CAGR), driven almost entirely by the release of ChatGPT, but this volume has not yet been matched by methodological depth: randomised controlled trials comparing LLM-integrated simulation with traditional approaches remain scarce.

Second, NTS research is heavily concentrated in communication (145 papers) and individual decision-making (135 papers), while the team-based competencies most critical to patient safety—teamwork, leadership, situational awareness, and crisis resource management—remain markedly underrepresented. Current LLM applications predominantly address one-to-one conversational tasks; the field urgently needs multi-agent systems capable of simulating team dynamics, CRM scenarios, and the real-time interpersonal demands of high-acuity clinical environments.

Third, virtual patient chatbots have emerged as the dominant simulation modality, yet their educational equivalence to human standardised patient encounters is unestablished. Mannequin-based and high-fidelity simulation, where LLM-driven voice interfaces could enhance realism and scalability, remains largely unexplored.

Fourth, urology simulation represents a near-complete gap, with only 6 papers identified and none examining LLM integration within established boot camp or NTS training formats—despite these programmes offering an ideal testbed for such technologies.

Fifth, the research ecosystem depends overwhelmingly on a single proprietary model (ChatGPT, 47% of papers), with open-source alternatives virtually absent. Systematic comparative evaluation across models is needed to ensure reproducibility and institutional autonomy.

Sixth, AI-assisted screening using independent LLM agents is a feasible and reliable approach to large-scale evidence synthesis, achieving agreement that exceeds published human-human benchmarks.

The convergence of LLMs and healthcare simulation has achieved remarkable breadth in fewer than four years but lacks the thematic balance, specialty-specific depth, and rigorous evaluation needed to guide the integration of these technologies into simulation-based NTS training with confidence.

## Data Availability

All data underlying the findings are available in the Supporting Information files and in the project repository at https://github.com/MattPears1/bibliometric-llm-healthcare-simulation. The submission version is available under the tag v1.0-submission at https://github.com/MattPears1/bibliometric-llm-healthcare-simulation/tree/v1.0-submission.

## SUPPORTING INFORMATION

Large Language Models in Healthcare Education and Simulation: A Bibliometric Analysis of 551 AI-Verified Papers (2020-2026)

Pears M, Wadhwa K, Payne SR, Konstantinidis ST, Biyani CS

## Appendix A: Search Strategies

Searches were conducted across seven open-access databases on 2026-03-22. A total of 35 query strings were executed, yielding 100,277 initial records. After deduplication (13,642 duplicates removed; 13.6% deduplication rate), 86,635 unique records remained for screening.

Search period: January 2020 to March 2026

Language restriction: English only

Proprietary databases excluded: Scopus, Web of Science (to maintain open-access reproducibility) Sequential Keyword Funnel (Stage 1):

- Step 1: LLM/AI filter (86,635 to 37,280 records) - Terms: "large language model," "LLM," "ChatGPT," "GPT-4," "Claude," "Gemini," "generative AI," "foundation model," and variants

- Step 2: Healthcare/medical/education filter (37,280 to 29,238 records) - Terms: "healthcare," "clinical," "medical education," "nursing," "patient," "training," and variants

- Step 3: Simulation filter (29,238 to 5,725 records) - Terms: "simulation," "virtual patient," "OSCE," "debriefing," "scenario," "mannequin," "VR," "XR," and variants

Stage 2: Tighter Second-Pass Filter (5,725 to 830 records):

Required: (a) named LLM; (b) medical/healthcare keyword in title; (c) education-related keyword; (d) exclusion of editorials/commentaries without original data.

Full query strings and keyword lists for all stages are available in the GitHub repository (Appendix F).

## Appendix B: PRISMA-trAIce Compliance Checklist

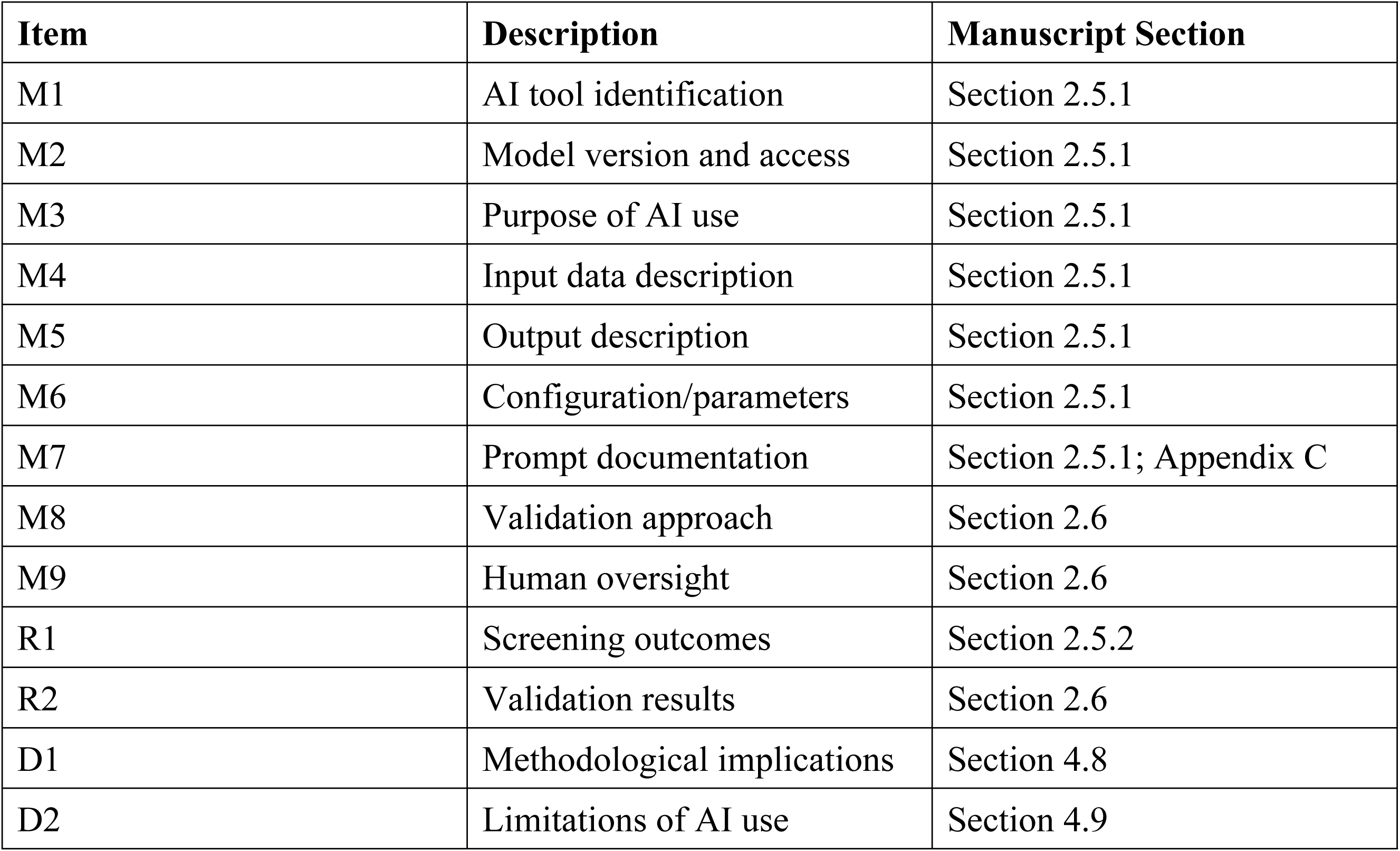

The following table maps each PRISMA-trAIce checklist item to the corresponding manuscript section.

## Appendix C: AI Screening Agent Prompt

The following prompt was given to each of 83 independent Claude Sonnet 4.6 agents (approximately 10 papers each):

INCLUSION CRITERIA - include if the paper addresses:

A) LLM/Generative AI as primary focus

B) Healthcare/medical context

C) Education or training context

EXCLUSION CRITERIA (exclude if ANY apply):

- Pure clinical AI with NO education element

- Non-healthcare context

- Traditional ML only (not LLM/generative AI)

- Editorial/commentary without original data

- LLM benchmarking without educational intervention TIER CLASSIFICATION:

- Tier 1: LLM + Healthcare Education (broadly)

- Tier 2: LLM + Healthcare Simulation

- Tier 3: LLM + Healthcare Simulation + Non-Technical Skills TAGS (true/false):

- simulation_relevant

- urology_relevant

- nts_relevant

For each paper, agents returned: binary include/exclude decision, confidence rating (high/medium/low), tier classification, sub-analysis tags, and a 1-3 sentence rationale.

Default API parameters were used. Each agent operated independently with no shared memory or cross-agent communication. The complete system prompt and per-paper instructions are available in the GitHub repository.

## Appendix D: Inter-Rater Reliability

The lead researcher (MP) independently reviewed 180 papers (21.7% of the 830-paper candidate corpus) across multiple validation stages.

Preliminary assessment (30 papers):

- Cohen’s kappa: 0.93 ("almost perfect")

- Confirmed sufficient agreement to proceed

Formal 100-paper assessment (random sample from 830-paper corpus):

- Pre-reconciliation agreement: 94%

- Pre-reconciliation Cohen’s kappa: 0.86 ("almost perfect")

- Sensitivity: 91.8%

- Disagreements: 6 (all exam benchmarking papers lacking educational intervention)

- Post-reconciliation Cohen’s kappa: 1.0 (perfect agreement)

- Post-reconciliation raw agreement: 100%

Both pre- and post-reconciliation kappa values exceed the mean human-human kappa of 0.82 for abstract screening reported by Hanegraaf et al. (2024).

Internal consistency review:

- All 830 agent-provided rationales examined against stated criteria

- Zero logical errors identified across all agents 50-paper pilot calibration:

- 64% inclusion rate on filtered corpus vs 22.5% on broader set

- Confirmed keyword filter enrichment of relevant papers

## Appendix E: Complete Figure Compendium

**Supplementary Figure S1:**
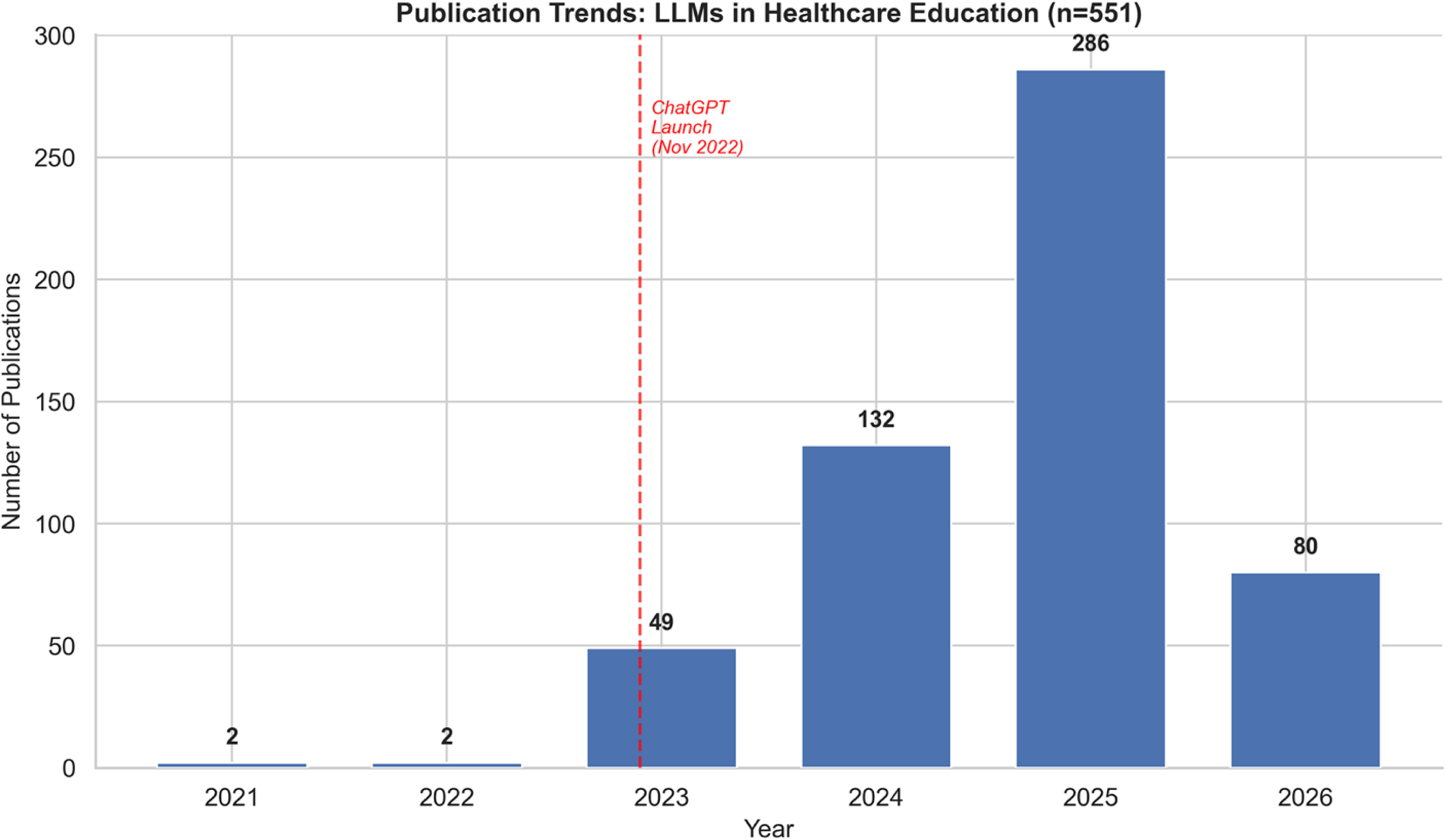
Publication Trends by Year.

Bar chart showing annual publication counts: 2021 (2), 2022 (2), 2023 (49), 2024 (132), 2025 (286), 2026 through March (80). Total: 551 papers. The ChatGPT launch (November 2022) is annotated. The compound annual growth rate is 109.1%.

**Supplementary Figure S2:**
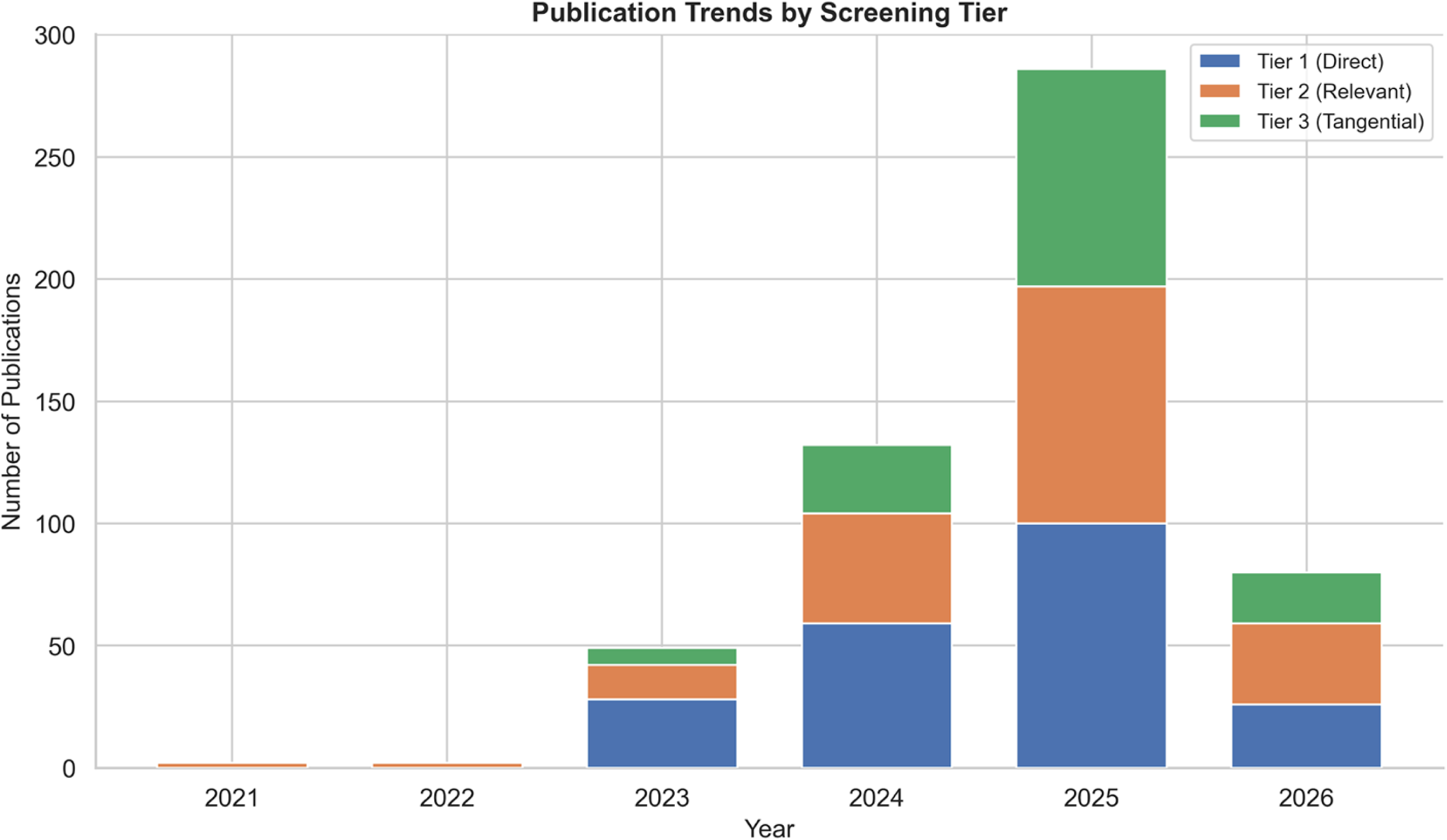
Publication Trends by Tier.

Stacked bar chart showing annual publication counts stratified by screening tier (Tier 1, Tier 2, Tier 3). All tiers show growth concentrated in 2024-2025, with Tier 3 (simulation + NTS) growing from 7 papers in 2023 to 89 in 2025.

**Supplementary Figure S3:**
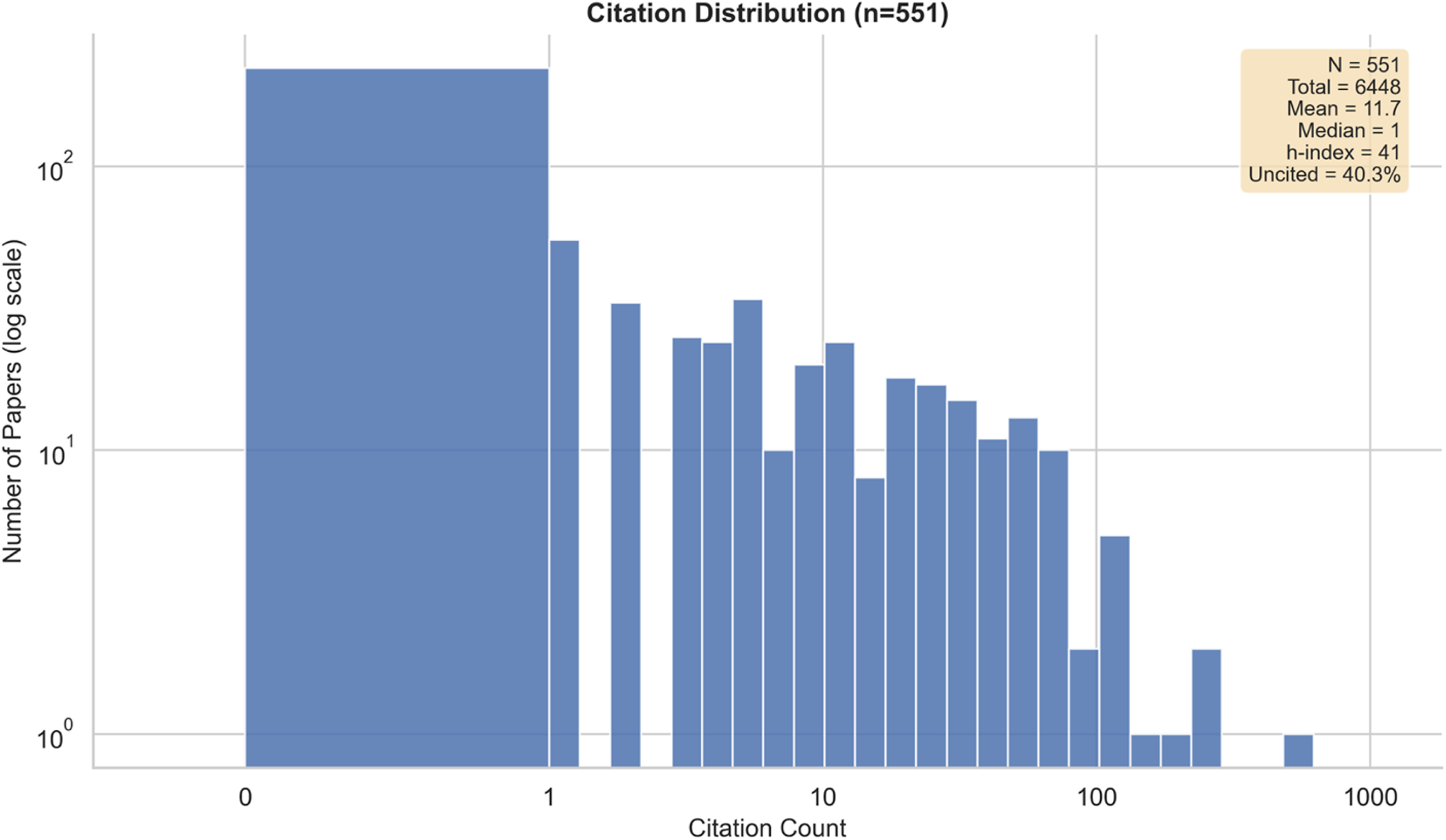
Citation Distribution.

Histogram of citation counts across the 551-paper corpus (log scale). Summary statistics: N = 551, total citations = 6,448, mean = 11.7, median = 1.0, h-index = 41, g-index = 69, i10-index = 131, uncited = 40.3% (222 papers). Most-cited paper: 550 citations.

**Supplementary Figure S4:**
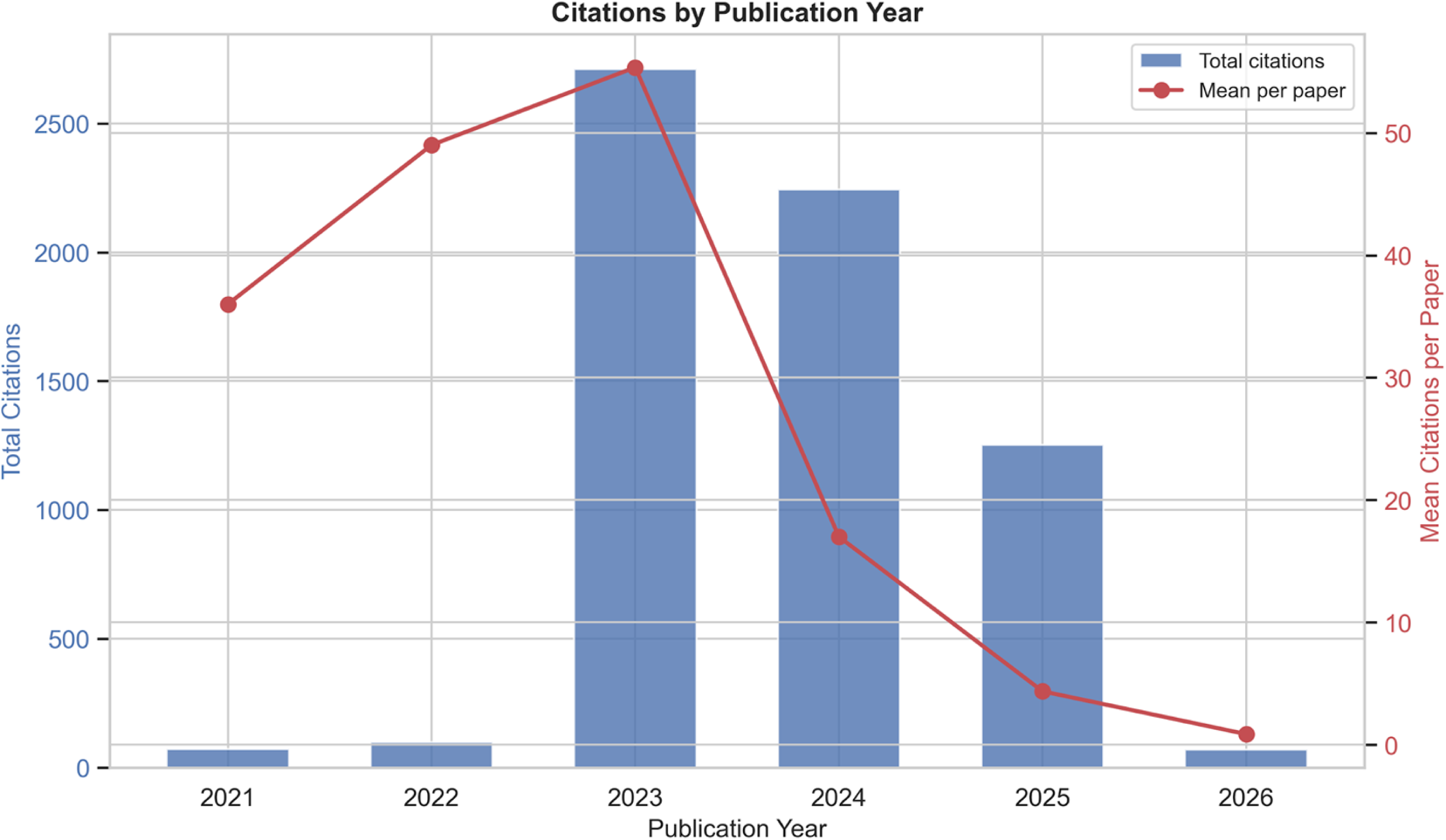
Citations Stratified by Year.

Dual-axis chart showing total citations per year (bars) and mean citations per paper (line). Mean citations per paper are highest for early publications (reflecting citation accumulation time) and decline for more recent years, while total citations peak in 2023.

**Supplementary Figure S5:**
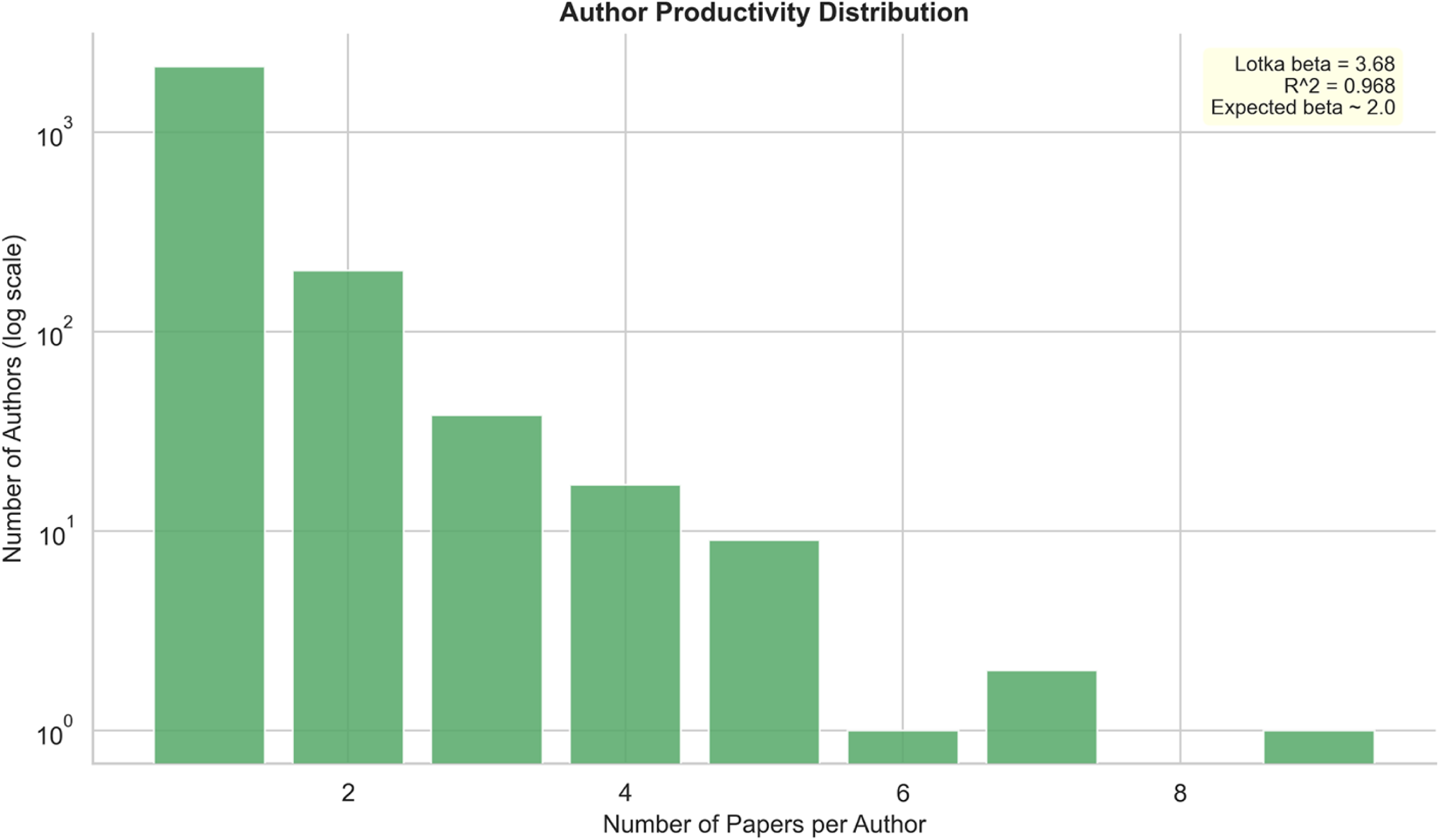
Author Productivity Distribution.

Log-scale distribution of author productivity. Of 2,398 unique authors, 2,128 (88.7%) contributed a single paper and 270 (11.3%) contributed two or more. Fitted Lotka exponent (beta) = 3.7 with R-squared = 0.97, exceeding the classical inverse-square value of 2.0.

**Supplementary Figure S6:**
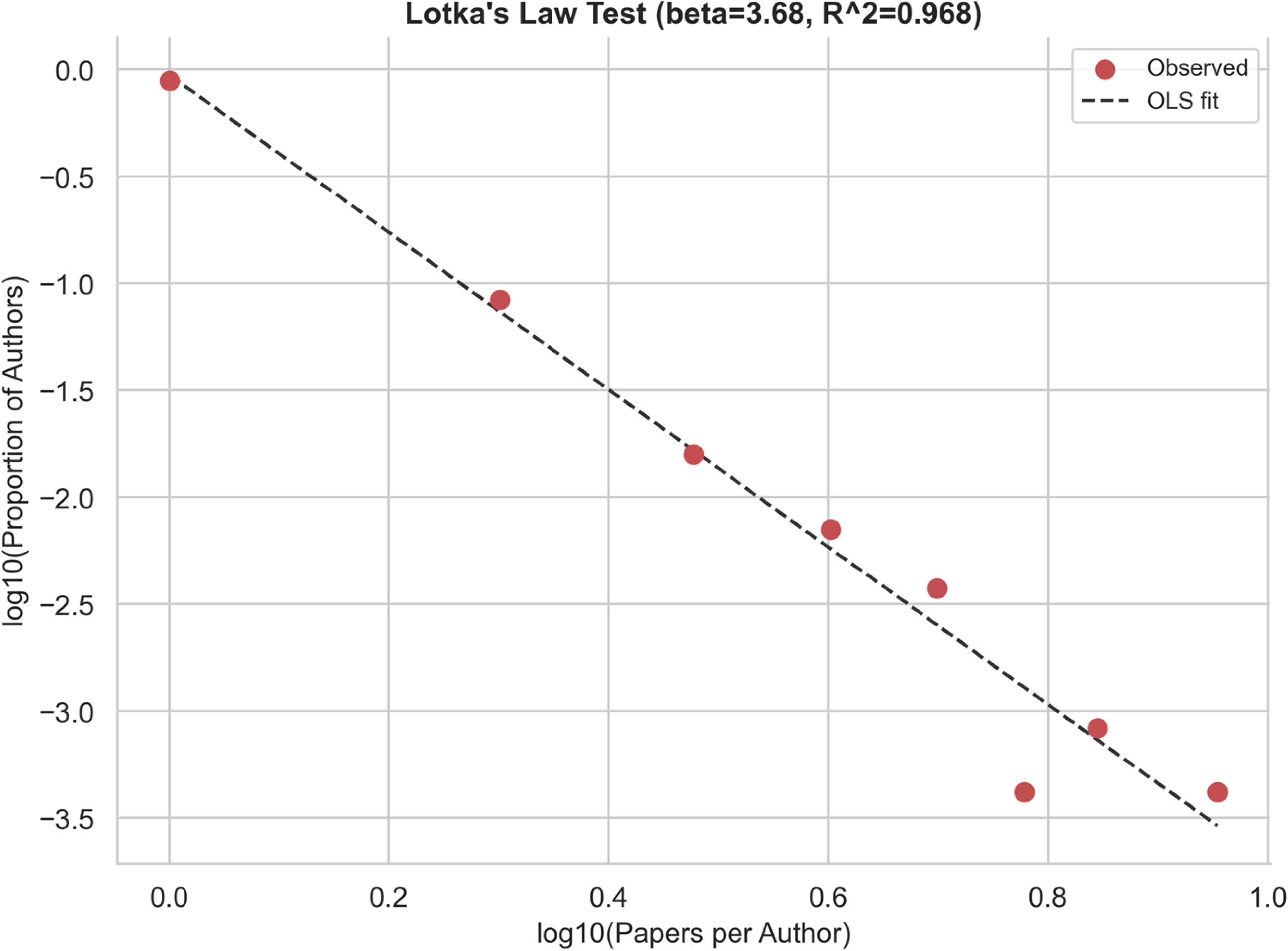
Lotka’s Law Fit.

Log-log plot of papers per author versus proportion of authors, with OLS regression line. Beta = 3.7, R-squared = 0.97, confirming a strong power-law relationship and highly skewed author productivity distribution.

**Supplementary Figure S7:**
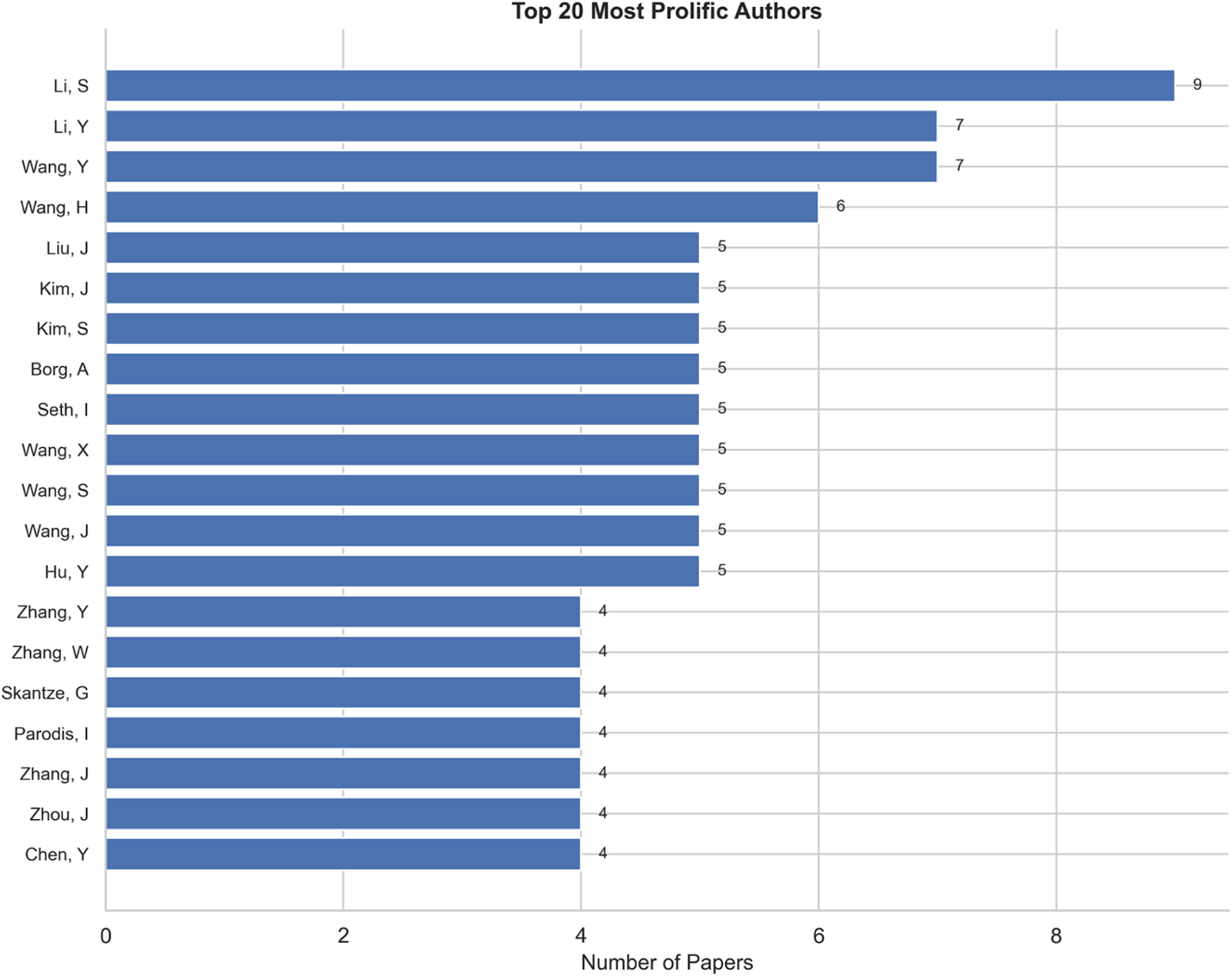
Top 20 Most Prolific Authors.

Horizontal bar chart of the 20 most productive authors. Li, S leads with 9 papers, followed by Li, Y (7), Wang, Y (7), and Wang, H (6). Note: common East Asian surnames may affect disambiguation.

**Supplementary Figure S8:**
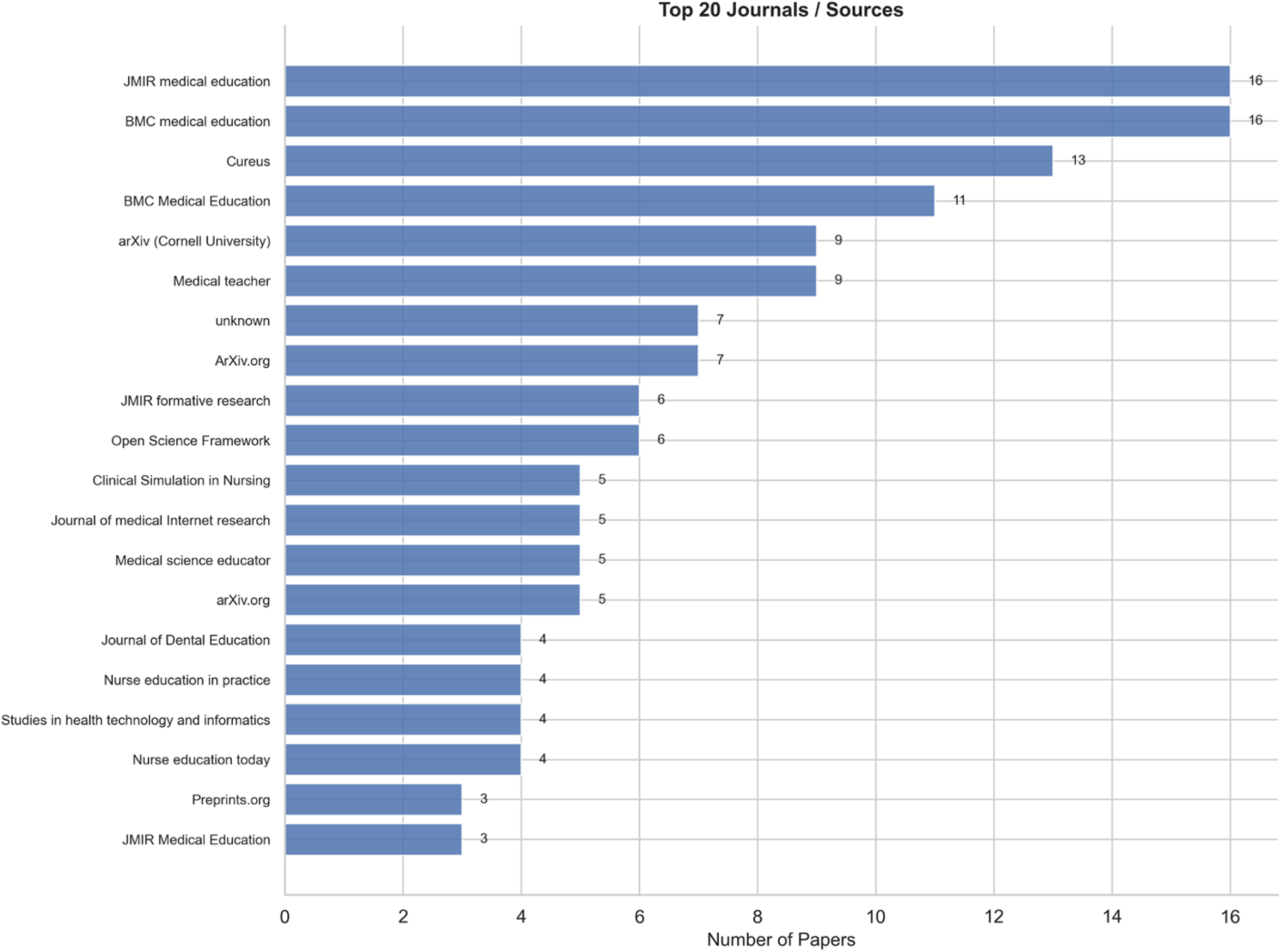
Top 20 Journals.

Horizontal bar chart of the 20 most productive journals. JMIR Medical Education and BMC Medical Education lead jointly with 16 papers each (3.7%), followed by Cureus with 13 papers (3.0%).

**Supplementary Figure S9:**
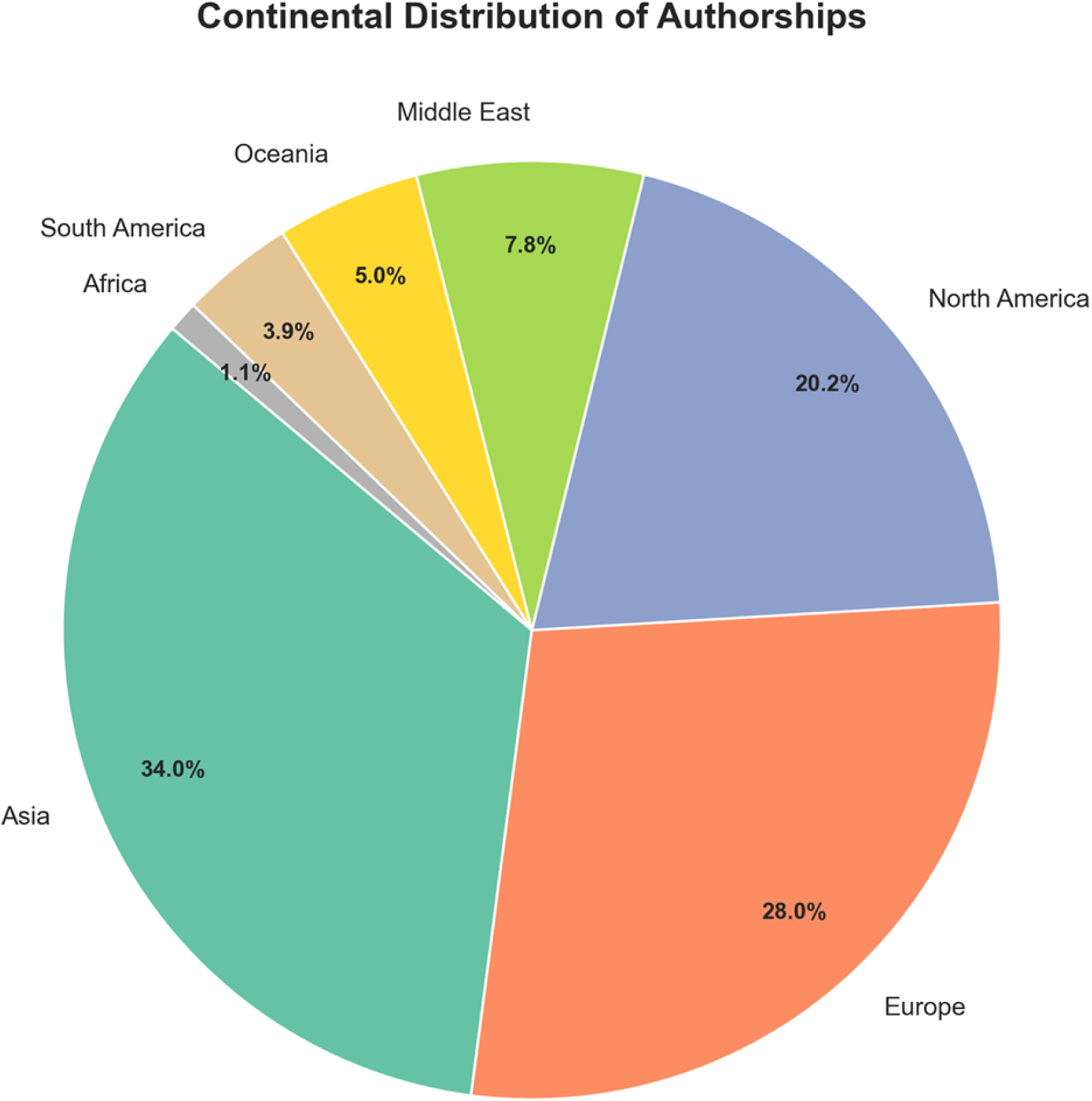
Continental Distribution of Publications.

Pie chart of authorship by continent (218 papers with geographic data): Asia (34.0%), Europe (28.0%), North America (20.2%), Middle East (7.8%), Oceania (5.0%), South America (3.9%), Africa (1.1%).

**Supplementary Figure S10:**
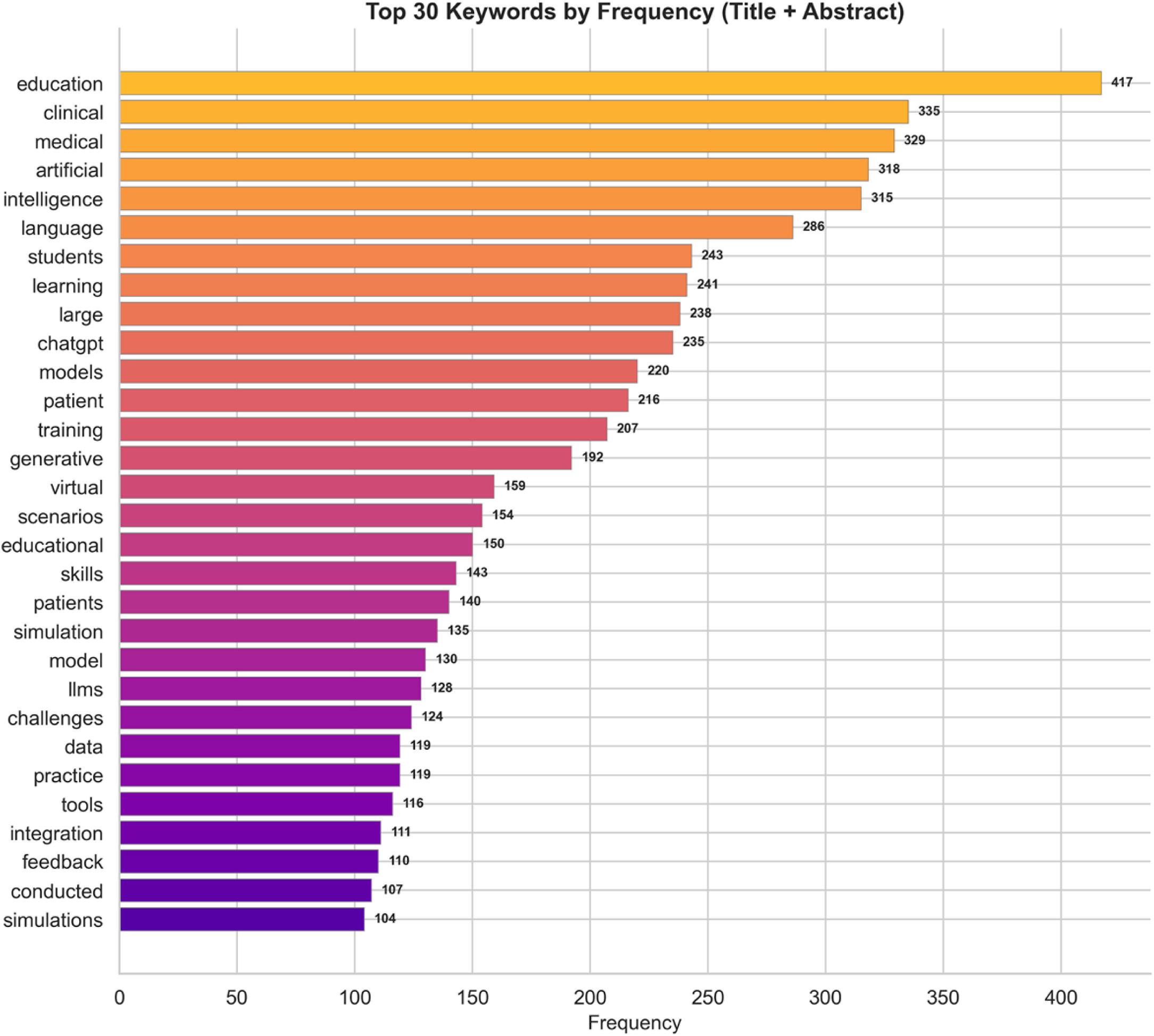
Top 30 Keywords.

Bar chart of the 30 most frequent keywords across the corpus. "Education" appears in 75.7% of papers, "clinical" in 60.8%, "medical" in 59.7%, "artificial" in 57.7%, and "intelligence" in 57.2%.

**Supplementary Figure S11:**
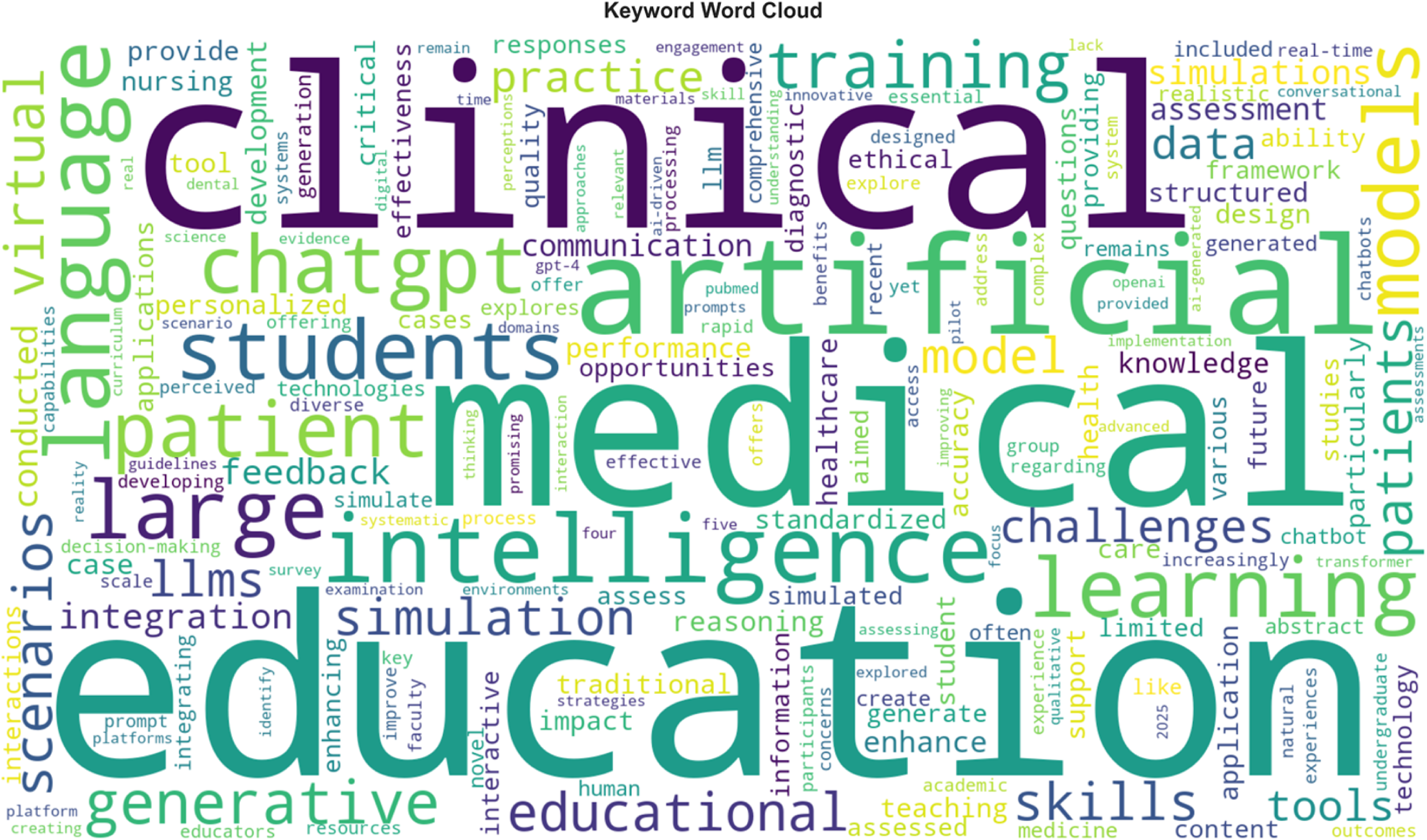
Keyword Word Cloud.

Word cloud visualisation of keyword frequencies, with word size proportional to occurrence count. Dominant terms include "education," "clinical," "medical," "artificial," "intelligence," and "chatgpt."

**Supplementary Figure S12:**
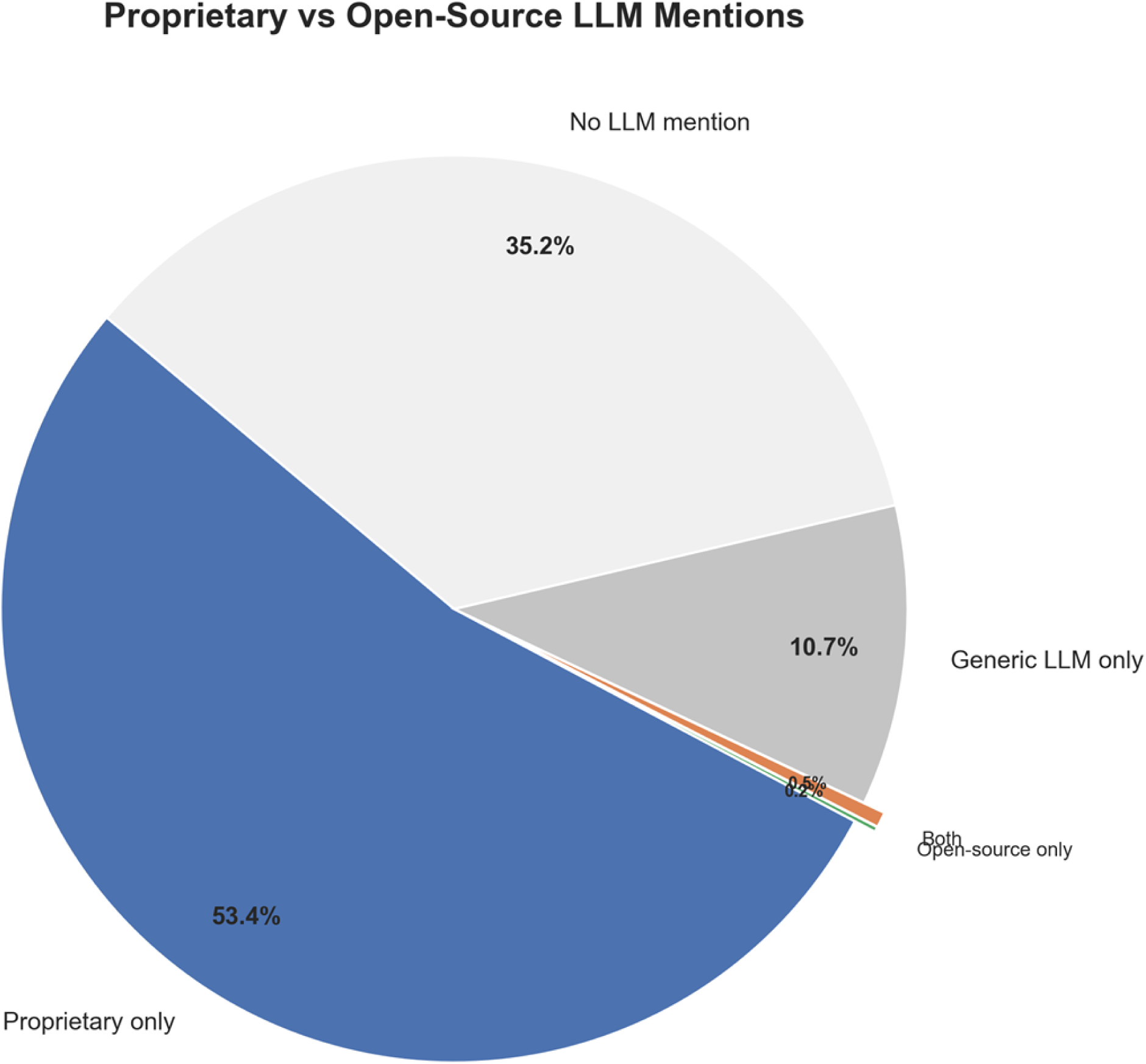
Proprietary vs Open-Source LLM Distribution.

Pie chart showing model licensing distribution: papers mentioning proprietary models only (53.4%), no specific model mentioned (remainder), open-source only (0.7%), and both proprietary and open-source (0.5%). Only 3 papers in the entire corpus compared proprietary and open-source models.

**Supplementary Figure S13:**
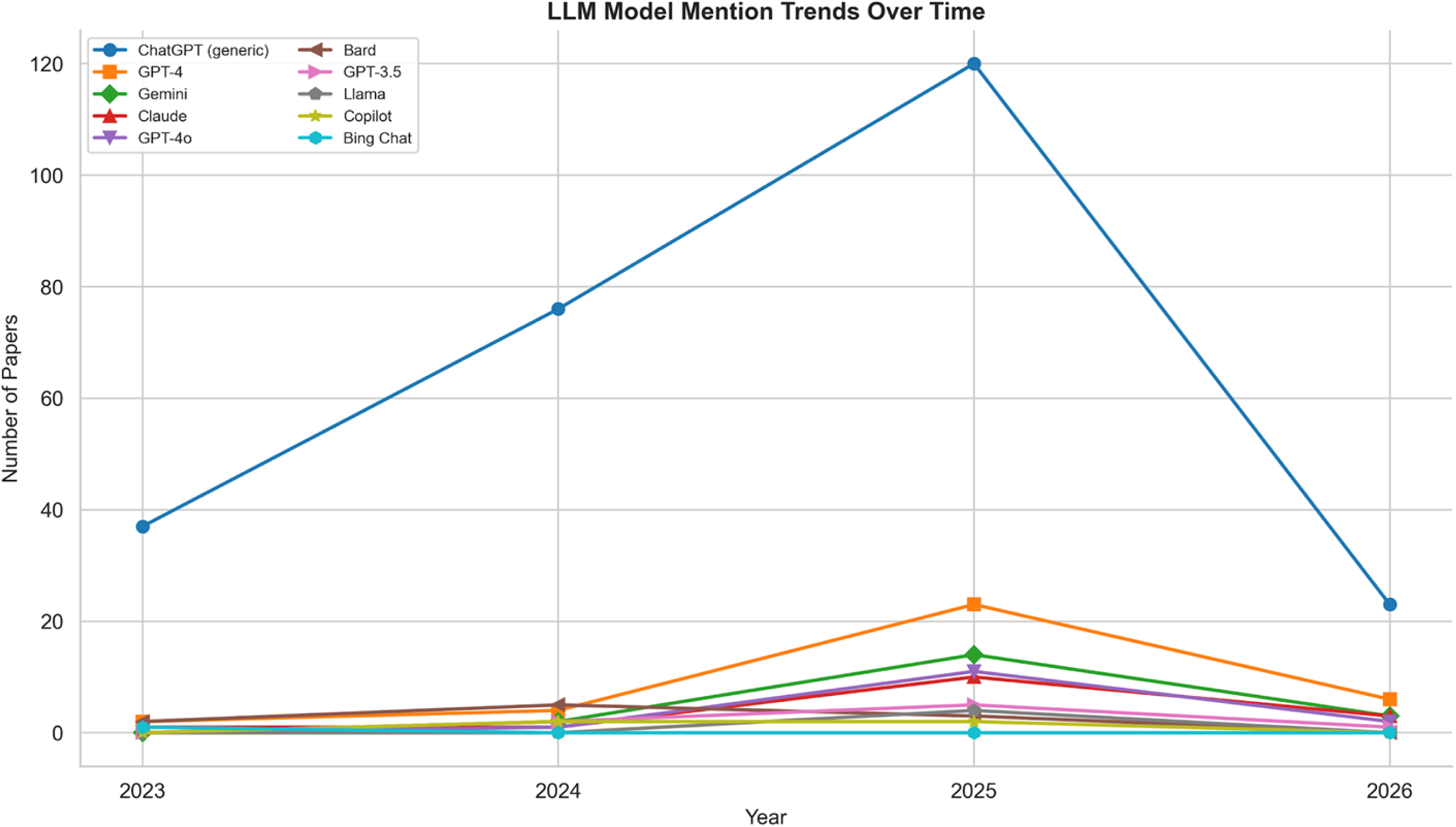
LLM Model Temporal Trends.

Line chart showing model mentions by year (2023-2026). ChatGPT dominates across all years. GPT-4 and Gemini emerge in 2024-2025. GPT-3.5 declines (model succession). Open-source models remain minimal throughout.

**Supplementary Figure S14:**
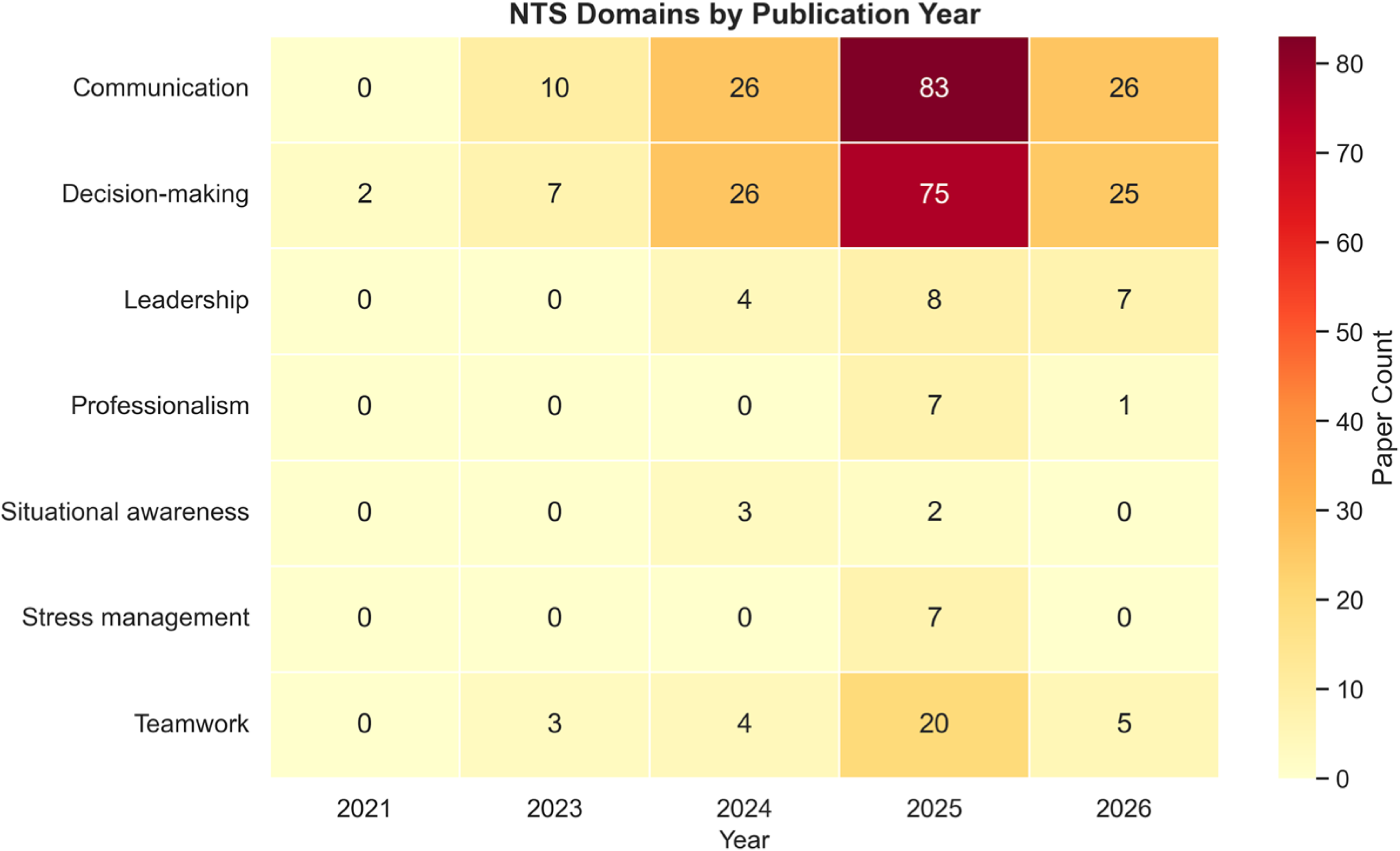
NTS Domain Temporal Heatmap.

Heatmap of NTS domain frequencies by year. Communication and Decision-making show the strongest growth (peaking in 2025 at 83 and 75 papers respectively). Teamwork, Leadership, and other domains remain low throughout. Year 2022 is absent (no NTS-relevant papers among the 2 total papers that year).

**Supplementary Figure S15:**
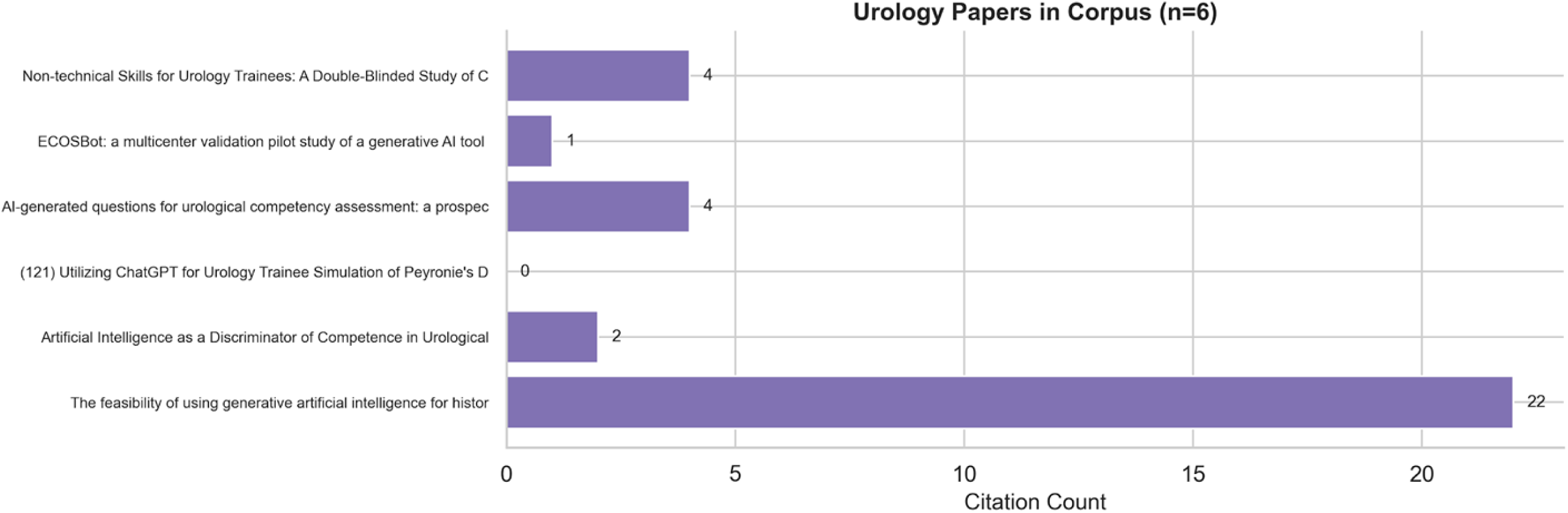
Urology Sub-Analysis.

Horizontal bar chart of the 6 urology-relevant papers with citation counts. All published in 2024-2025. Total citations: 33, mean: 5.5. NTS domains represented: Communication (2 papers), Decision-making (1 paper). No papers address teamwork, leadership, situational awareness, or CRM in urology.

**Supplementary Figure S16:**
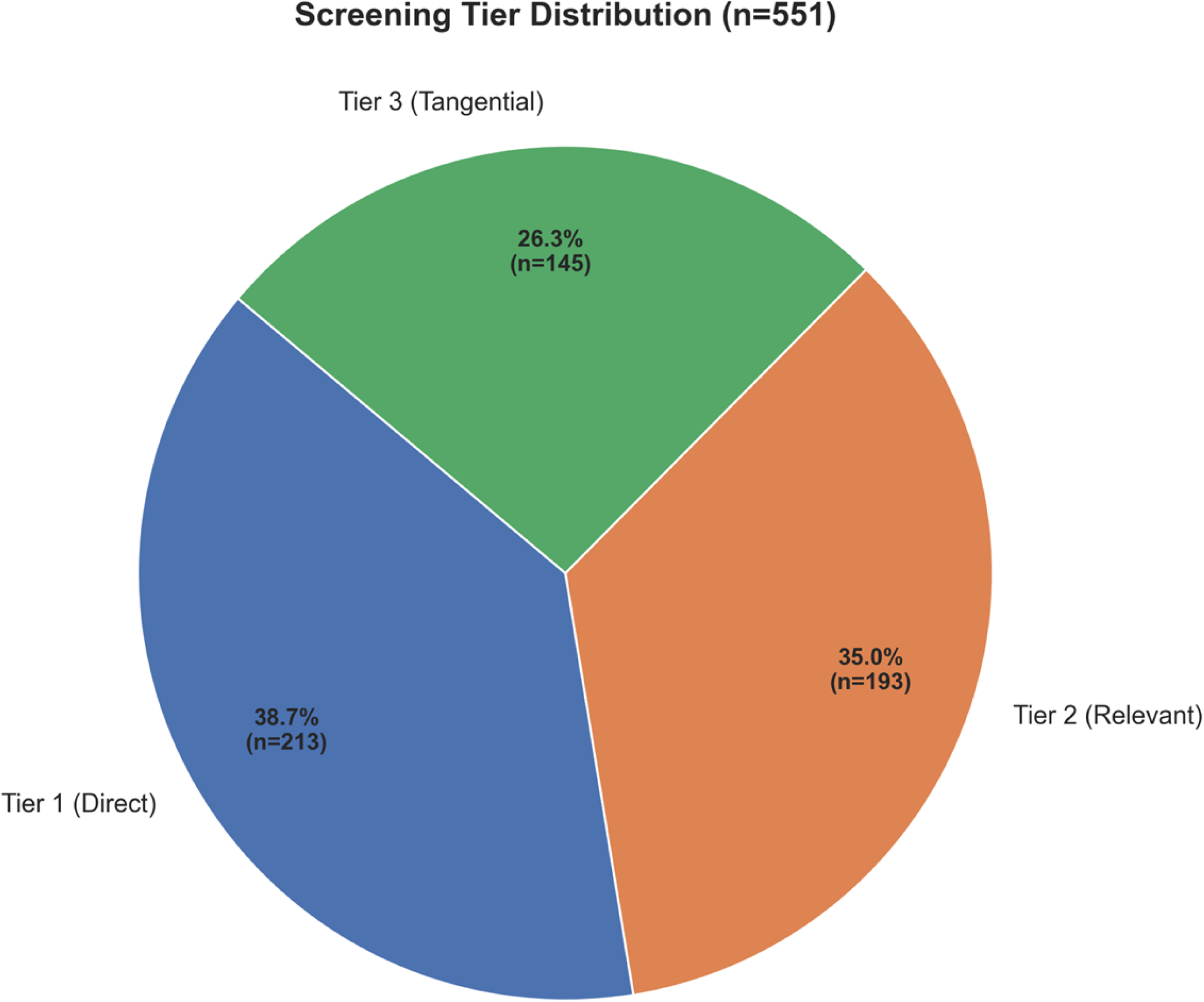
Tier Distribution.

Pie chart showing the three-tier corpus composition: Tier 1 - LLMs in healthcare education broadly (213 papers, 38.7%), Tier 2 - LLMs in healthcare simulation (193 papers, 35.0%), Tier 3 - LLMs in healthcare simulation with NTS (145 papers, 26.3%).

**Supplementary Figure S17:**
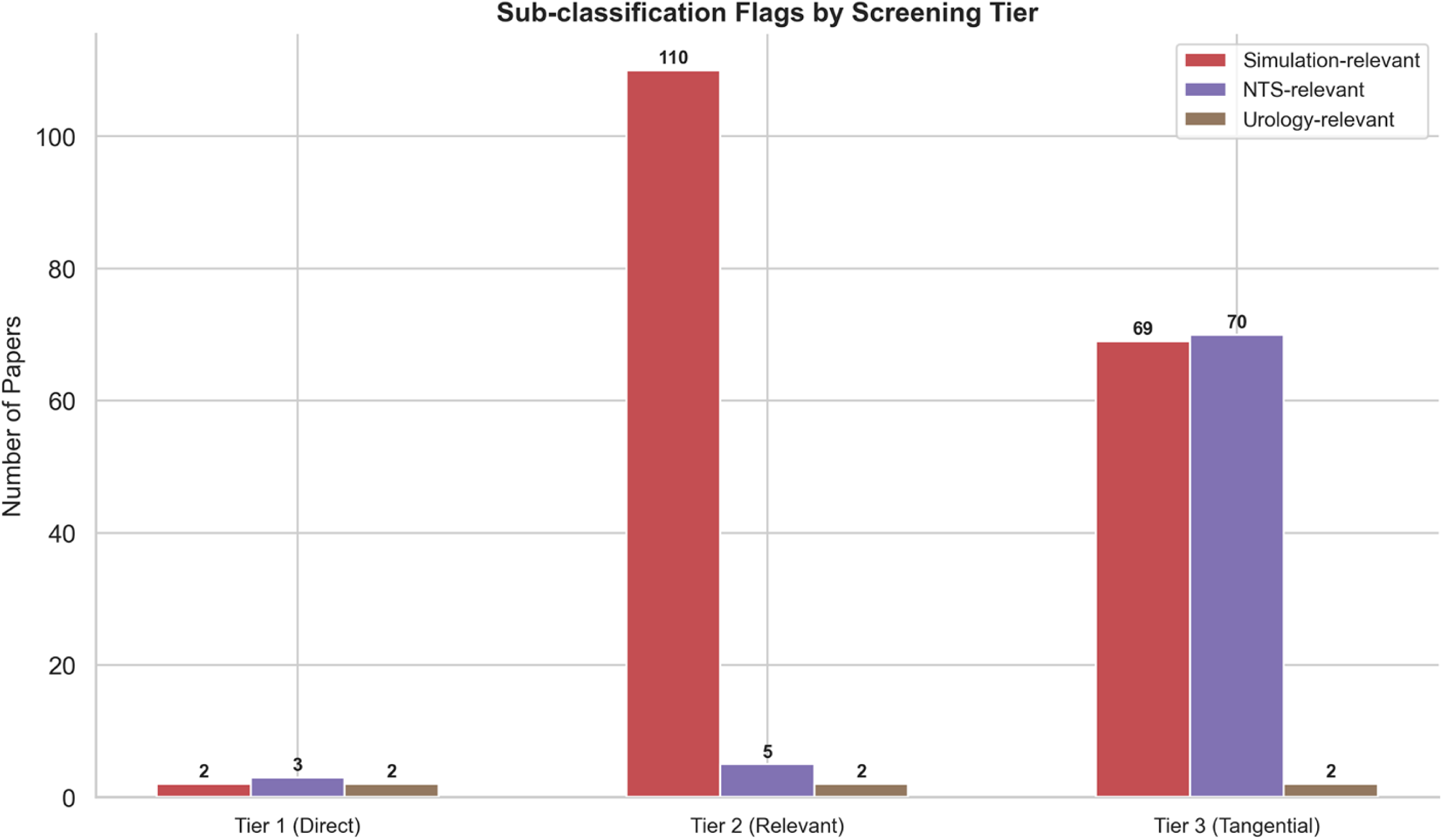
Sub-Analysis Flags by Tier.

Grouped bar chart showing the distribution of simulation-relevant, NTS-relevant, and urology-relevant flags across tiers. Tier 2 has the highest simulation relevance (110 papers); Tier 3 has the highest NTS relevance (70 papers). Urology is evenly distributed (2 per tier).

**Supplementary Figure S18:**
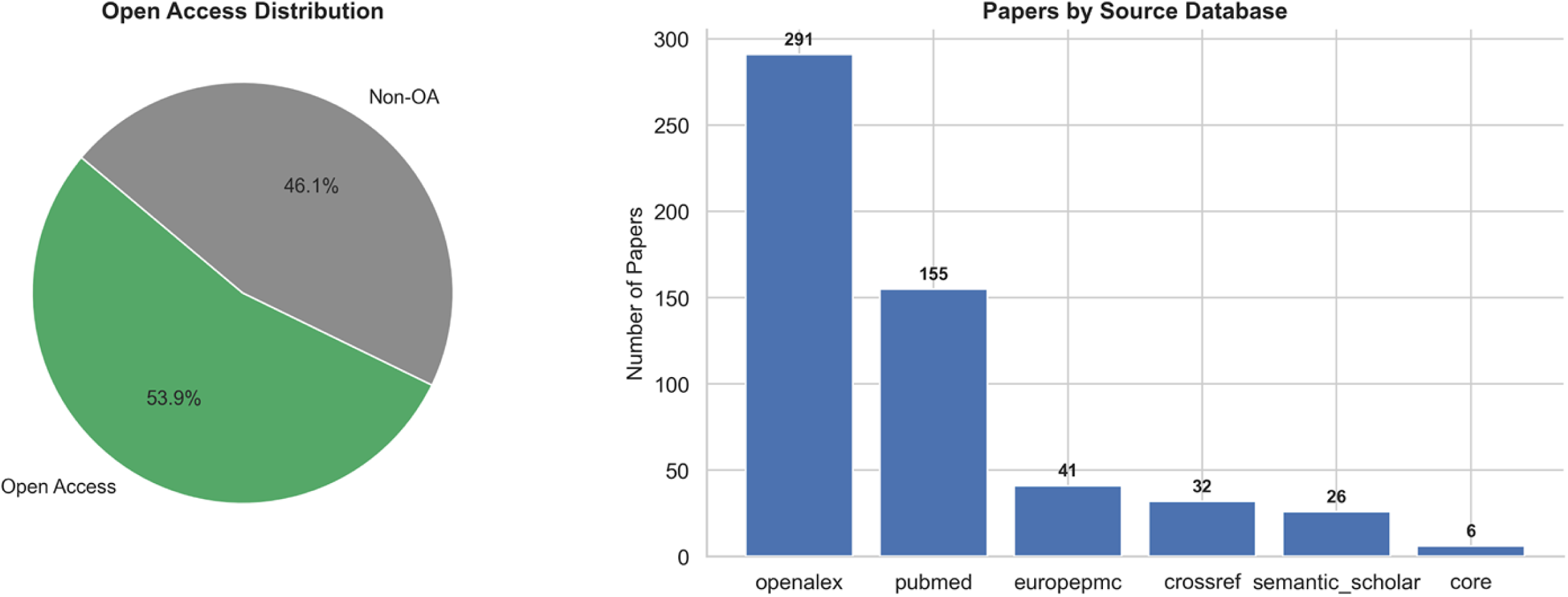
Descriptive Overview Dashboard.

Summary dashboard showing open-access distribution (297 papers, 53.9% open access) and database source breakdown: OpenAlex (291), PubMed (155), Europe PMC (41), Crossref (32), Semantic Scholar (26), CORE (6), DOAJ (0).

## Dataset Summary

| **Metric** | **Value** |
| --- | --- |
| Total papers (AI-verified corpus) | 551 |
| Initial records retrieved | 100,277 |
| Databases queried | 7 (all open-access) |
| Search date | 2026-03-22 |
| Year range | 2021-2026 |
| Language | English only |
| Open access papers | 297 (54%) |
| AI screening tool | Claude Sonnet 4.6 (83 agents) |
| Human validation | 180 papers (22% of candidates) |
| Cohen's kappa | 0.86-1.0 |
| CAGR | 109% |
| h-index | 41 |
| Total citations | 6,448 |
| Unique authors | 2,398 |
| Countries | 58 |
| Journals | 279 |
| Keywords | 6,412 |

Initial records retrieved: 100,277

Databases queried: 7 (all open-access)

Search date: 2026-03-22

Year range: 2021-2026

Languages: English only

Open access papers: 297 (53.9%)

AI screening tool: Claude Sonnet 4.6 (83 independent agents)

Human validation: 180 papers (21.7% of candidate corpus)

Inter-rater reliability: Cohen’s kappa 0.86-1.0

Key metrics:

- CAGR: 109.1%

- h-index: 41

- Total citations: 6,448

- Unique authors: 2,398

- Countries represented: 58

- Unique journals: 279

- Unique keywords: 6,412

## Appendix F: Code and Data Availability

Repository: https://github.com/MattPears1/bibliometric-llm-healthcare-simulation

The repository includes:

- Data collection scripts for all 7 databases

- Deduplication and preprocessing pipeline

- Sequential keyword filtering implementation

- AI screening agent deployment code

- Complete system prompts and per-paper instructions

- Raw agent outputs with rationales for all 830 screened papers

- Bibliometric analysis scripts (publication trends, citation analysis, author productivity, geographic analysis, keyword co-occurrence, LLM model classification, simulation modality classification, NTS domain classification, urology sub-analysis)

- Figure generation scripts

- Core corpus dataset (551 papers with metadata)

Requirements: Python 3.13+

This work was funded by The Urology Foundation Innovation and Research Award 2025.

The funder had no role in study design, data collection and analysis, decision to publish, or preparation of the manuscript.

---

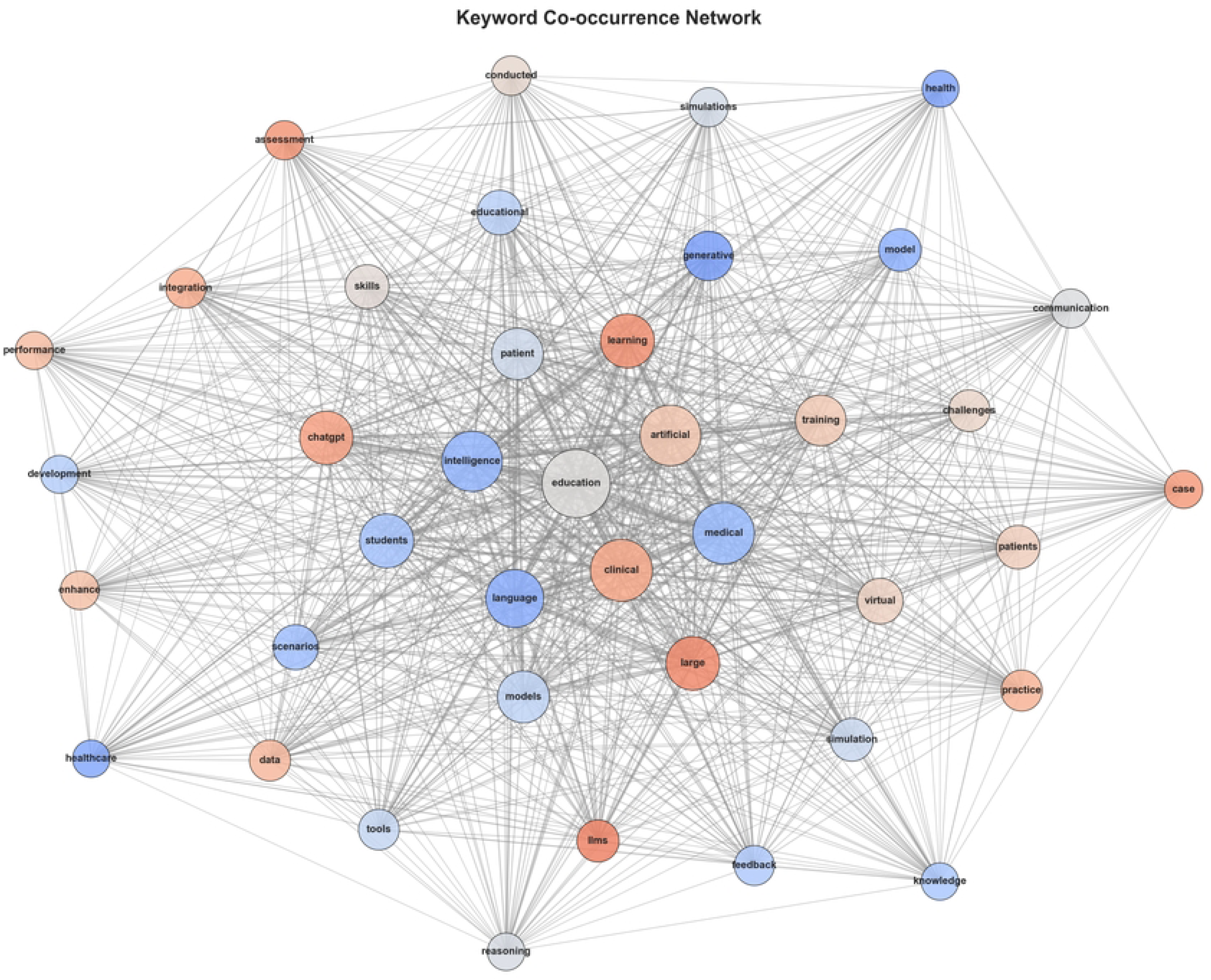

